# Extreme temperature exposure and the risk of miscarriages in Italy: a nationwide small-area study of 4.5 million births

**DOI:** 10.64898/2026.07.02.26356686

**Authors:** Sandra Gudžiūnaitė, Emiliano Ceccarelli, Jane Hirst, Monica Pirani, Alice Maraschini, Hanns Moshammer, Giada Minelli, Marta Blangiardo

## Abstract

**Background:** The effect of extreme temperatures on miscarriage is not well understood. Even less understood is the gestational period most vulnerable to extreme temperature exposure, as early miscarriages are often missed in incident datasets. We employ a birth–rate based approach to infer the risk of miscarriage in response to extreme temperature exposure by gestational week.

**Methods:** We conducted a population-based ecological study using birth registry data from the 7,948 municipalities of Italy between 2013 and 2024 (4.5 million births). The analyses were stratified by five climatically coherent macro–regions (Ecoregions). To infer unreported pregnancy losses, we regressed birth rates dated from the last menstrual period against weekly temperatures across gestational weeks 3–21, accounting for temporal seasonality and spatial heterogeneities.

**Findings:** Exposure to heat (mean weekly temperature of 30 · 4 *^◦^*C) during gestational weeks 3-4 was associated with a reduction of birth rates of 1.62 (0.71 - 2.51) –%, and of 1.91 (0.92 - 2.88) –% to mean weekly temperature of 1 · 6 *^◦^*C. Whilst heat was found to be harmful during gestational weeks 3-4 and 18-21, cold spells were found to be consistently harmful from the 3^th^ up to the 12^th^ week, depending on the Ecoregion.

**Interpretation:** Pregnancies are vulnerable to extreme temperatures during the post-conceptual period and the second trimester. The findings underscore the need for a pre-conceptual cohort to clarify the mechanisms of loss, and urge public health action to protect pregnancies from the beginning of gestation.

## 1 Introduction

Gestation is marked by a sequence of critical developmental milestones, which can be disrupted by adverse environmental exposures, and result in congenital malformation, pregnancy loss, or restricted growth [1]. For instance, organogenesis, occurring between the 5^th^ and 9^th^ gestational weeks (GW), is considered to be an important sensitive period, in which the developing fetus is most sensitive to teratogens [2].

The gestational periods during which pregnancies are most vulnerable to miscarriage following exposure to extreme heat or cold remain poorly understood.

A core obstacle to studying miscarriage is the lack of comprehensive, population-level data sources. Birth registries are a commonly used data source for studying birth outcomes; however, they systematically exclude miscarriages as they only capture pregnancies that reach the gestational threshold for viability, usually around the 20^th^ GW, in which a fetus is considered to be able to survive outside the womb [3]. Hospital records present a different but related limitation, since they only capture losses that required clinical intervention, missing miscarriages that occur in the earliest gestational weeks, often before pregnant women are even aware of the pregnancy [4]. Similarly, in cohort studies, the degree of left-censoring depends on when the observation window opens and how follow-up is conducted, meaning that very early losses are frequently missed regardless of study design. It is uncommon for the observation period to begin at the point of intention to conceive, since women tend to learn of their pregnancies in the 6^th^ week of gestation [5], thus a substantial proportion of early losses may occur before a pregnancy is ever recorded, contributing further to under-reporting of incidence.

Italy presents a particularly relevant context for studying the perinatal health effects of extreme temperatures. As a Mediterranean country with diverse morphology and climatic zones [6], Italy’s temperature plummet during autumn and winter and remains cooler through the summer in the mountains, whilst some territories experience intensely hot summers, with heatwaves becoming increasingly frequent and severe [7], posing a growing public health concern. Epidemiological evidence from Italy links gestational heat and cold exposure to increased risks of preterm birth and low birth weight [8], but the evidence on miscarriages remains scarce, with only a handful of studies globally.

In this study we use a Bayesian spatiotemporal model to evaluate the impact of extreme temperatures on miscarriages over 10 years across the 7,948 municipalities in Italy. Using a similar approach to Kioumourtzoglou et al. [9], we use the birth registry to reconstruct the number of weekly conceptions and link this to the temperature over the post-conceptual gestational weeks before viability (22 gestational weeks). This allows us to estimate the gestational weeks most vulnerable to extreme temperatures as well as to obtain the cumulative effect of extreme temperature across the entire period. Our aims are to assess whether early gestational exposure to extreme temperatures displays evidence of miscarriages in the population, and to identify the gestational weeks that are most vulnerable to extreme exposures.

### 1.1 Methods

#### 1.1.1 Study design

This is a population-based ecological time series study where we used municipality level data on weekly births to estimate the dose–response relationship between temperature across gestational weeks 3–21 and birth rates. We infer miscarriages associated with transient exposure to heat or cold by a reduction of birth rates. The gestational period of analysis captures the post-conceptual period, up to the pregnancy viability threshold after which births — live or dead — are captured in the Italian birth registry [3].

#### 1.1.2 Study area

Italy is a Mediterranean peninsula in Southern Europe, extending southward from the Alpine mountain chain in the north along the Apennines mountain range, which forms the country’s geographical backbone. The Adriatic Sea borders Italy to the East, while the Tyrrhenian Sea lies to the West, off which Italy’s two largest islands - Sicily and Sardinia - are situated. As of 2025, Italy’s population stood at 58, 942, 828 [10], distributed across 7, 948 municipalities.

The models were stratified spatially according to the municipalities’ classification into Ecoregions, large territorial units, geographically distinct, with a similar composition and distribution of ecosystem resources [6]. This classification follows a hierarchical approach, grouping municipalities into territorial units with increasing internal homogeneity based on climatic, biogeographical, physiographic, and hydrographic factors into 5 ecoregions: *Alpine Chain*, *Po Valley*, *Apennine Chain*, *Tyrrhenian and the Islands*, *Apulia and the Adriatic Coast*. A map of Italy by Ecoregion is presented in the Supplementary Figure A.1.

#### 1.1.3 Birth data and inference of pregnancy loss

The outcome data were sourced from the Italian Birth Registry “Certificato di Assistenza al Parto” (CeDAP) [11]. We limited the analysis to singletons born between years 2013 and 2024 of gestational ages 22–42. We only included births from women aged 16 to 49, and we excluded those whose municipality of birth was not reported.

For each birth, the last menstrual period (LMP) week was approximated by subtracting the gestational age in weeks from the date of birth. We then aggregated births by LMP week and maternal municipality of residence. As offset we include in the analysis the female population aged 16–49 by municipality, obtained as yearly counts from the Italian National Institute of Statistics (Istituto Nazionale di Statistica, ISTAT). We interpolated the yearly estimates linearly to obtain weekly values.

#### 1.1.4 Meteorological Data

We extracted ambient temperatures at two meters above surface level from the ERA5-Land reanalysis [12], a fifth-generation gridded reanalysis produced by the European Centre for Medium-Range Weather Forecasts (ECMWF), which provided hourly grids at 9×9k^°^m resolution. These were processed further into weekly averages to be used as the main exposure in the analysis. We allocated values to each municipality by computing the weighted spatial averages of the grids falling within a municipality’s geometry, as detailed in Section A.2 of the Supplementary Materials.

#### 1.1.5 Confounders

Our analyses were adjusted for nitrogen dioxide (NO_2_) levels, a proxy of traffic related air pollution. Data was extracted from the Copernicus Atmosphere Monitoring Service (CAMS), a gridded satellite-based reanalysis product [13], providing hourly grids at 10×10 km resolution, which we processed further into weekly averages. We used the same approach as with the temperature to assign NO_2_ to each municipality (see A.2 of the Supplementary Materials). To account for socio-economic confounding, we considered the employment rate of the population aged 20 to 64, extracted from the composite “Fragility Index” (Indice di Fragilità Comunale) last updated in 2021 [14].

### 1.2 Statistical model

The analysis was conducted in a Bayesian hierarchical framework, stratified by Ecoregion. Birth counts were assigned a Negative Binomial distribution due to overdispersion in the data. The weekly mean temperature was assigned at the municipality level and linked to each birth traced to GW 3–21. To capture potentially non-linear and delayed effects of temperature across early pregnancy, we modelled both the intensity of exposure and its timing using flexible functions that allow for non-linear and lagged associations through a distributed lag non-linear model (DLNM). We included the employment rate as a fixed effect, assuming a linear relationship with the outcome. We incorporated spatial random effects to account for unmeasured heterogeneity between municipalities as well as neighbourhood-based spatial dependency [15]. The temporal component was decomposed into a long-term trend, modelled as a linear fixed effect for year, and a seasonal component, captured by Fourier sine-cosine terms. Lastly, a combination of residual structured and unstructured spatial effect was included in the model.

Sensitivity analyses were conducted to assess the robustness of findings to alternative modelling assumptions, including different specifications of seasonal patterns, exclusion of preterm births, and alternative functional forms for the temperature-response relationship. A more detailed overview of the methods, including the model specification and the sensitivity analyses is provided in the Supplementary Materials D.1.

Results are presented as relative risks (RR) of birth rates. A RR below 1, indicating a reduction in birth rates, observed in association with extreme temperature exposure (defined as temperatures at the 90th and the 10th percentiles relative to the median temperature of each Ecoregion-specific distribution) during a given gestational window (weeks 3–21) is interpreted as evidence of increased pregnancy loss associated with that exposure. We report the GW-specific effects, as well as then sum the GW-specific RR across the post-conceptual period (GW 3-4) and the period of organogenesis (GW 5-9). The gestational period in which the association is strongest is interpreted to be the one in which the pregnancy is most sensitive to extreme temperatures. This inference rests on the assumption that, after accounting for seasonality, spatial heterogeneity, and other confounders, deviations in birth rates during the pre-viability gestational period reflect changes in pregnancy loss rates, following the approach established in [9] for traffic air pollution exposure.

We provide the interpretation of the model parameters in terms of relative risks, e.g. a change in the birth rates upon exposure to an extreme temperature (10th or 90th percentile of Ecoregion-specific distribution) specific temperature at given gestational period is compared with a reference (here the median or the 10% and 90% when focusing on extreme cold and warm periods).

### 1.3 Implementation and software

All analyses were conducted using R [version 4.4.2, 16]. Bayesian models were estimated using r-INLA [17], while cross-basis functions were constructed using the dlnm package [18]. The code to replicate the analyses is available on GitHub at https://github.com/sanderulis/inferring_miscarriages_italy.git. The area-level exposure levels from gridded datasets were assigned using the terra package.

## 2 Results

### 2.1 Descriptive analysis

A total of 4, 457, 468 singleton births of gestational weeks 22–42, of which 4,204,116 (94%) full term, were included in the 11-year study period, with first week of gestation starting between 2013-01-07 and 2023-12-25. A detailed study profile and attrition table is reported in Table B.1.1 of the Supplementary Materials. Figure C.1 of the Supplementary Materials reports the trend in the daily number of births from LMP stratified by Ecoregion, showing a decreasing trend across Italy, and marked seasonality of births. Daily mean temperatures showed a clear increasing trend over time across all Italian Ecoregions (Fig. C.2 of the Supplementary Materials). A marked north–south gradient was also evident, with lower values recorded in the Alpine Chain (mean 12·96*^◦^*C, standard deviation 8·37*^◦^*C) and progressively higher values towards southern and coastal areas, particularly in the Apulia and Adriatic ecoregion (mean 20·10*^◦^*C, s.d. 7·70*^◦^*C).

### 2.2 Model results

#### 2.2.1 Spatial and temporal birth rate patterns

Figure 2.1 shows the estimated seasonal pattern of the birth rates across the Ecoregions for the period under study. Generally a strong seasonality is visible, with LMPs resulting in highest birth rates falling in the months of December and January and a dip between May and July; however weaker seasonal patterns are seen in the Po Valley and the Alpine Chain Ecoregions.

**Figure 2.1:**
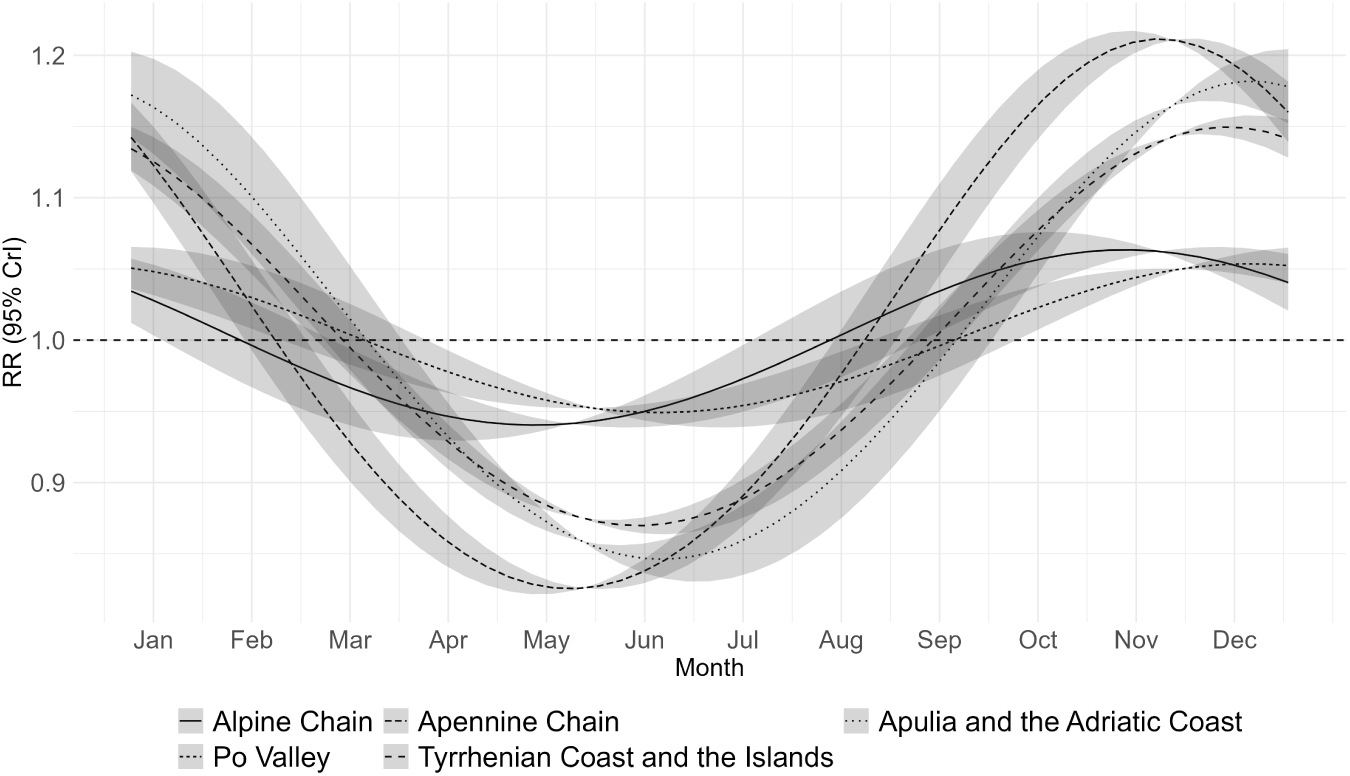
Seasonal yearly component of birth rates from LMP week.

The spatial variability of birth rates is plotted in the Supplementary Figures C.5. Generally birth rates are lower in the north-central regions, particularly in Tuscany in the West; the Southern regions show small clusters of very high values, but at the same time are characterised by large variability, particularly in the two main islands of Sicily and Sardinia.

### 2.3 Temperature and birth rate relationship

In what follows, reductions in birth rates associated with extreme temperature exposure during the post-conceptual, pre-viability gestational period (GW 3–22) are interpreted as evidence of increased pregnancy loss, as described in the Statistical Model section.

The association between overall temperature exposure throughout the GWs 3–21 and birth rate is displayed in Figure 2.2, and reported in the Supplementary Table 6. Taking the median temperature of each Ecoregion as reference, we see a detrimental effect of cold spells throughout the period of interest on birth rates in all regions except for the Po Valley, with estimated birth rates decreasing as much as 9·05% (CrI 95% 11·69 – 6·33) for temperature at the 10^th^ percentile of the all-year distribution (−3·1 C*^◦^*) compared to the median (12·5 C*^◦^*) in the Apennine Chain, whereas exposure to the 90^th^ percentile (28·1C*^◦^*) throughout GWs 3–21 corresponded to relative increase of birth rates of 16·37% (CrI 95% 8·6 - 23·8). The 10^th^ and 90^th^ percentiles of the Ecoregion-specific distribution are reported in Supplementary Table B.3.

**Figure 2.2:**
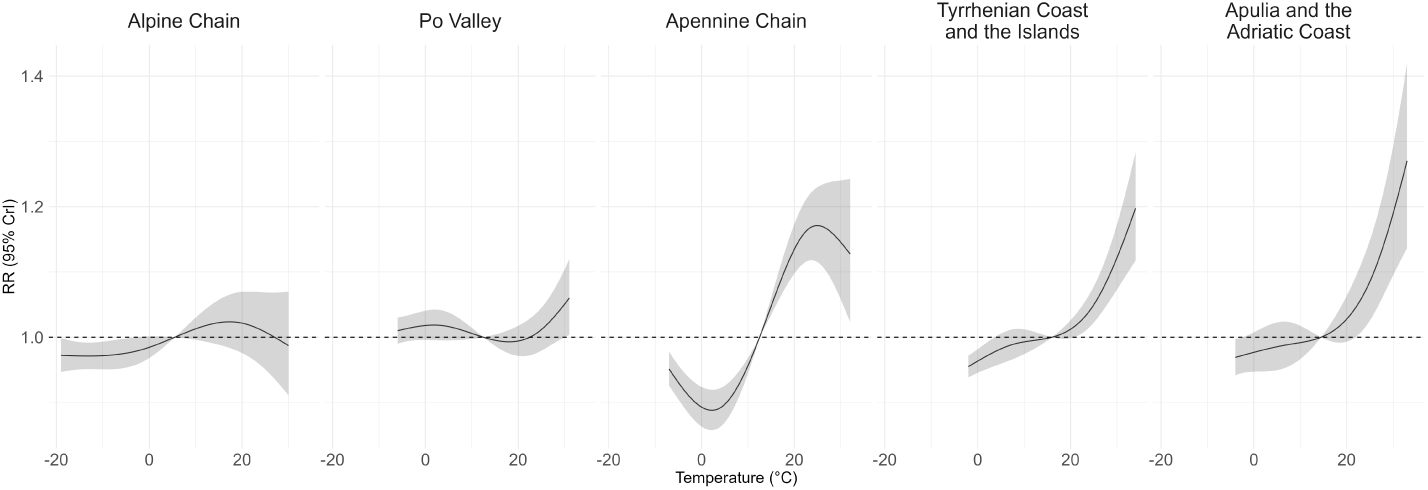
Overall Association between mean temperatures during GW 3–21 and birth rates.

We then focused on the effects of exposure during specific subsections of the gestational periods. The post–conceptual period was defined across GWs 3–4, Figure 2.3. Throughout it, the association between temperature and birth rates appears to follow an inverted–U shape across all Ecoregions, suggesting that extreme cold and warm temperatures are detrimental over the first few weeks of the pregnancy. The strongest association was found in the Po Valley, where cold exposure resulted in a drop of 1·54% (2·34 – 0·74), and heat of 1·70% (0·82 – 2·52) relative to the median temperature.

**Figure 2.3:**
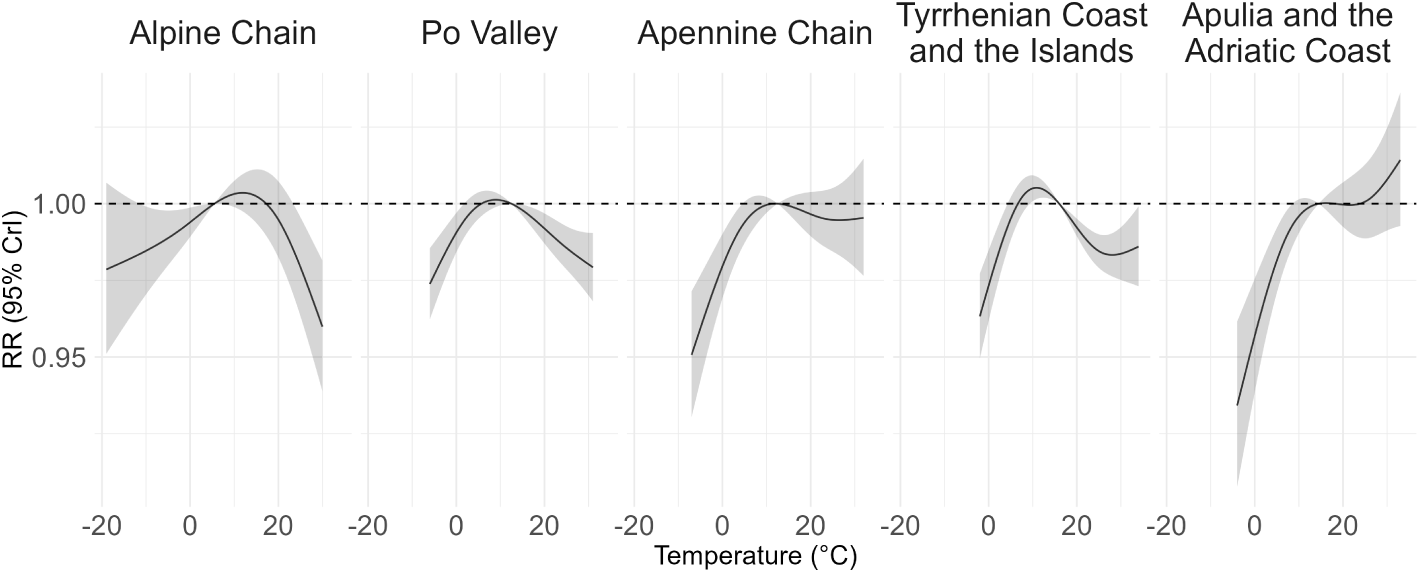
Association between temperature over GW 3–4.

**Figure 2.4:**
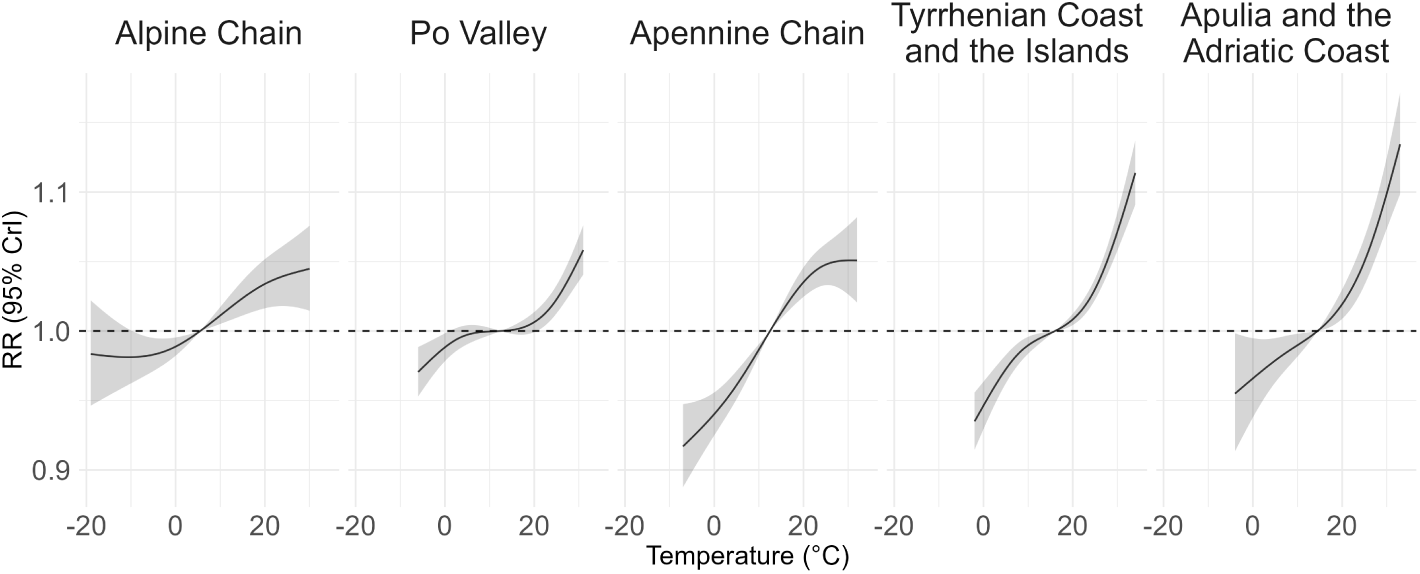
Association between temperature over GW 5–9.

The Appennine and the Alpine region shows large uncertainty for temperature below 0, while the Appennine Chain and the Apulia and Adriatic Coast Ecoregions are characterised by high uncertainty for the warm temperatures, likely due to data sparsity. The temperature percentile specific associations are reported in Supplementary Figure 9.

In the organogenic period, spanning between GW 5–9, the model shows a harmful effect only of cold temperatures across all regions, and a protective effect of heat, estimating reduction in the birth rates down to −1·80% (CrI 95% −0·55 – −3·04) for an average weekly temperature of −2·4 C*^◦^*in the Po Valley compared to the median (12·5), and an increase by 3·5% (CrI 95% 2·30 – 4·71) upon heat exposure (27·3 *^◦^*C) 2.4. The temperature percentile specific associations are reported in Supplementary Figure 10.

Since the impact of extreme temperatures varies across gestation, we present the GW–specific birth rate response to temperatures at the 90^th^ and the 10^th^ Ecoregion–specific percentile, to identify the most vulnerable windows. The GW–specific exposure to heat relative to the median presents an inverted-U-shape across all Ecoregions, with exposure throughout the 3–5^th^ and the 18–21^th^ gestational weeks associated with lower birth rates across the five Ecoregions.

The strongest association is found in the Tyrrhenian Coast and the Islands, where exposure to a mean weekly temperature of 30 · 4*^◦^*C during the 4^th^ GW corresponds to a drop by 1·19% (CrI 95% 1·68 – 0·69), and in the 19^th^ gestational week, by 0·38 % (CrI 95% 0·05 – 0·70).

The GW–specific exposure to cold appears to be detrimental on birth rates only in the first few weeks after conception, but shows a degree of variability across Ecoregion: whilst the detrimental effect appears to be smaller and only last up to the 7^th^ GW in the Po Valley, with a reduction of birth rates by 0.18 (CrI 95% 0·02 – 0·34) in the 6^th^ GW, whereas in the Alpine Chain this corresponded to a drop by 0·77 (CrI 95% 0·07 – 0·28)), whereas Apulia and the Adriatic have the strongest but less precise effect. No Ecoregion showed a second dip in the second trimester.

**Figure 2.5:**
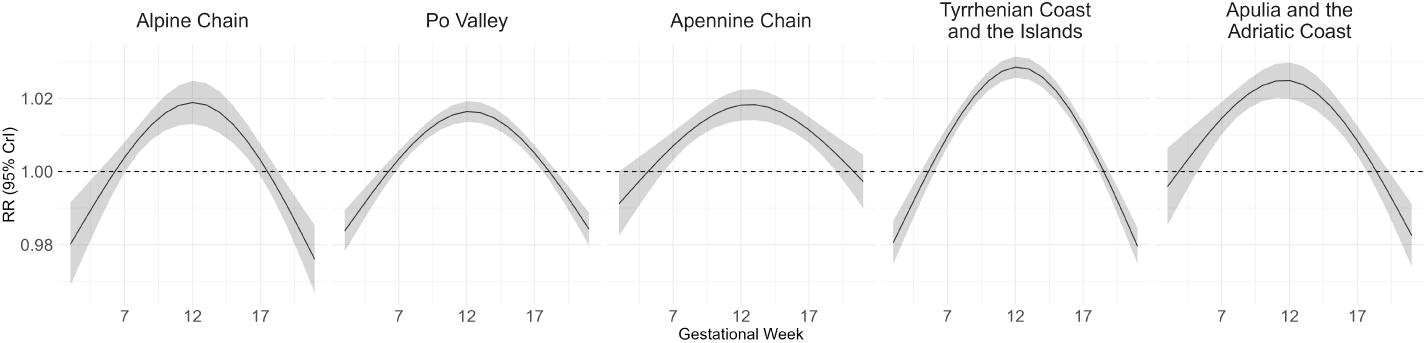
Gestational week-specific association between exposure to temperatures above the ecoregion specific 90th percentile of mean temperatures.

**Figure 2.6:**
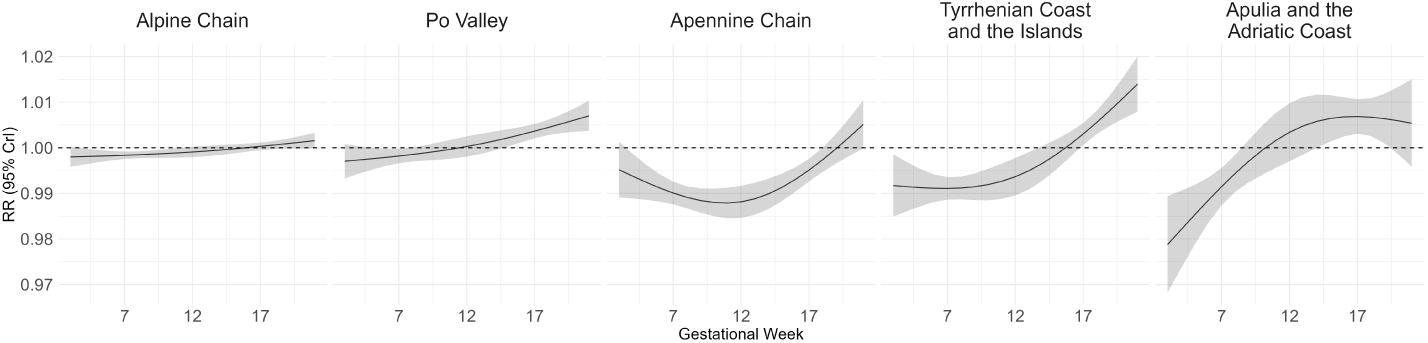
Gestational week-specific association between exposure to temperatures at the Ecoregion specific 10th percentile of mean temperatures.

## 3 Discussion

This is a nation-wide, spatiotemporal overview of birth rates in Italy’s 7, 948 municipalities over the period 2013–2024 investigating whether post-conceptual exposure to extreme temperatures is associated with miscarriages. We found that exposure to extreme heat, defined as average daily temperatures above the Ecoregion 90th percentile, during the immediate post conceptual period (GW 3–4) and the 4th month of gestation (GW 18–21) is associated with an increase risk of pre-viability pregnancy losses. This finding was consistent across all Ecoregions. Conversely, extreme cold exposure is associated with a detrimental effect on viable pregnancies in the early post-conceptual period (up to the 5^th^ GW in the Tyrrhenian and the Islands). To our knowledge, this is the first study to report on the burden of extreme temperature exposures on miscarriages in Italy, and one of the few nationwide studies to have been conducted worldwide. Although the effects of gestational exposure to extreme temperatures on miscarriage risk is understudied [19], a small number of epidemiological publications supports the association between heat and miscarriages. Wesselink and colleagues [20] examined incident miscarriage events, reporting a non-significant increase in miscarriage odds following exposure to extremely hot days (OR: 1 · 2, 95% CI: 0 · 95 − _°_1 · 5) and no association with cold exposure. The absence of a cold effect in that study may be partly explained by administrative data limitations: if cold exposure is most impactful in early gestation, studies relying on administratively recorded incident data — which typically suffer from high left-censoring gestational ages — are unlikely to capture this early vulnerability window. Overall, there is a meagre body of research looking at sensitive periods to heat exposure on miscarriages, and inconsistencies on how sensitive windows are operationalised. Hajdu at al. [21] used a similar approach to our study, and also found that exposure to hot temperatures during the first few weeks post-conception increases the clinically unobserved pregnancy loss rate, and exposure to colder temperatures seems to decrease it. In contrast, Shah et al. [22] found that in Nicosia, exposure to both higher and lower temperatures in the early pregnancy (GW 1–9) increased miscarriage risk. Our findings of heat and cold sensitivity at the immediate post-conceptual period is concordant with both studies. We additionally identify a sensitive period to heat in the second trimester, a finding, to the best of our knowledge, not previously reported in miscarriage studies.

Our results lend empirical support to proposed mechanistic pathways through which extreme temperatures may disrupt early post-conceptual processes. Nevertheless, evidence from humans remains scarce, and the underlying mechanisms are largely inferred from animal models or extrapolated from downstream physiological processes. Heat exposure may disrupt progesterone activity [23], thereby impairing uterine preparation for implantation. Additionally, heat may trigger inflammation-mediated fetal rejection through the upregulation of Heat Shock Protein 70 (HSP70) [24], and reduce blood flow to the placenta, thereby increasing the risk of pregnancy loss in the organ-growth phase of gestation (12GW+). Mechanisms for cold include immune activation, which may disrupt the delicate inflammatory balance required for successful implantation and early placental invasion [25]. These mechanisms are consistent with the post–implantation sensitive period to cold identified in this study.

A birth rate based approach to estimate the burden of missed pregnancies has been used in econometrics, and was first proposed to estimate miscarriage burden in response to traffic-related air pollution [9]. A key strength of this design is its resistance to under-reporting, a limitation shared by the vast majority of studies relying on incident miscarriage data. This means we can assess the burden of extreme temperature exposures on very early gestation, before the usual notification of pregnancy (typically before GW 6 [26]), capturing losses that would be missed in incident datasets.

A further advantage is that the approach yields gestational-week specific effect estimates, enabling identification of the most vulnerable windows of gestation. We were able to explore possible adaptations to temperature by the meaningful spatial stratification of the analysis into Ecoregions, which partition the 7, 984 municipalities into morphologically and climatically coherent units, which reflect Italy’s diverse temperatures, elevation, and coastal influences.

The validity of our approach depends on seasonal and geographic variation being adequately captured by the model. Seasonality modelled via Fourier sine-cosine terms may not fully account for irregular or complex annual patterns, although sensitivity analyses with alternative seasonal specifications were conducted to assess the robustness of findings to this assumption. Similarly, while the spatial random effect accounts for both spatially structured and unstructured heterogeneity, it may not fully resolve fine-scale geographic variation arising from localised phenomena such as urban heat islands, leaving residual spatial confounding possible. Furthermore, the LMP, and thus the GW-specific exposure, is back-calculated from the gestational age at birth. This variable is subject to error stemming from inaccurate maternal recall, or inconsistent clinical estimation. Any misclassification would propagate directly into the exposure assignment, potentially distorting estimated associations across gestational weeks.

These findings, together with the largely concordant body of literature, underscore the importance of developing specific policy levers aimed at protecting pregnant people, as well as those trying to conceive. Looking ahead, the burden of extreme heat exposure is projected to worsen considerably, with heatwave-attributable mortality expected to increase by an average of 140% annually [27]. Italian response to the climate crisis is formalised in the National Adaptation Plan for Climate Change (PNACC), which recognises elderly, children and those in lower socioeconomic groups amongst the most vulnerable populations [28], which does not make explicit statements about pregnant women. The clinical management of pregnancy is primarily coordinated by the Italian National Institute of Health (Istituto Superiore di Sanità – ISS) [29], however, their guidelines currently lack specific recommendations or protocols regarding the clinical management of pregnancy during heatwaves, and mitigation strategies are left to be implemented by local authorities. Our findings underscore the particular vulnerability of the immediate post-conceptual period. Yet social norms around early pregnancy disclosure, combined with limited public awareness of the risks faced during the peri-conceptual period, mean that pregnant women are least likely to receive adequate protection precisely when they are most at risk.

To conclude, this study suggests that both extreme cold and heat exposures increase miscarriages, with pregnancies most vulnerable to heat during the immediate post-conceptual period and the mid-second trimester, and to cold during the first weeks after conception. These findings call for the inclusion of pregnant women as well as those trying to conceive in national climate adaptation plans, and highlights the need for further research into sensitive gestational windows of vulnerability to extreme temperature, ideally through a pre-conceptual prospective cohort designed to capture the effects of transient extreme temperature exposures on the earliest gestational period.

## Data Availability

Data from the Italian Birth Registrys Certificate of Childbirth Assistance (CeDAP) are available from the Istituto Superiore di Sanita (ISS), but access to them is subject to restrictions; these data were used under license for the purposes of this study and are therefore not publicly accessible. However, the data are available from the authors upon receipt of a justified request and subject to authorization by the ISS. Requests for access to the CeDaP dataset should be sent to.

## Acknowledgements

S.G. is part of the the Doctortal Training Programme “Science and Solutions for a Changing Planet” Scholarship (SSCP-DTP) provided by the Natural Environment Research Council (NERC) through the Grantham Institute for Climate Change and the Environment.

E.C. was supported by the European Union – NextGenerationEU, under the PNRR project “C_PA - DM118 P.A. Pubblica Amministrazione: Metodologie statistiche per il supporto alle decisioni in contesto sanitario pubblico: stima dell’impatto degli eventi climatici estremi sulla salute della popolazione generale e la costruzione di modelli di allerta rapida”, CUP: B53C23002660006, Missione 4, Componente 1 (Missione I.4.1, PNRR Scholarships for Public Administration).

J.E.H. is supported by a UKRI Future Leaders Fellowship (MR/Y033833/1)

M.B and M.P A acknowledge infrastructure support for the Department of Epidemiology and Biostatistics provided by the NIHR Imperial Biomedical Research Centre (BRC).

## A Supplementary Methods

### A.1 Italy by Ecoregions

**Figure A.1:**
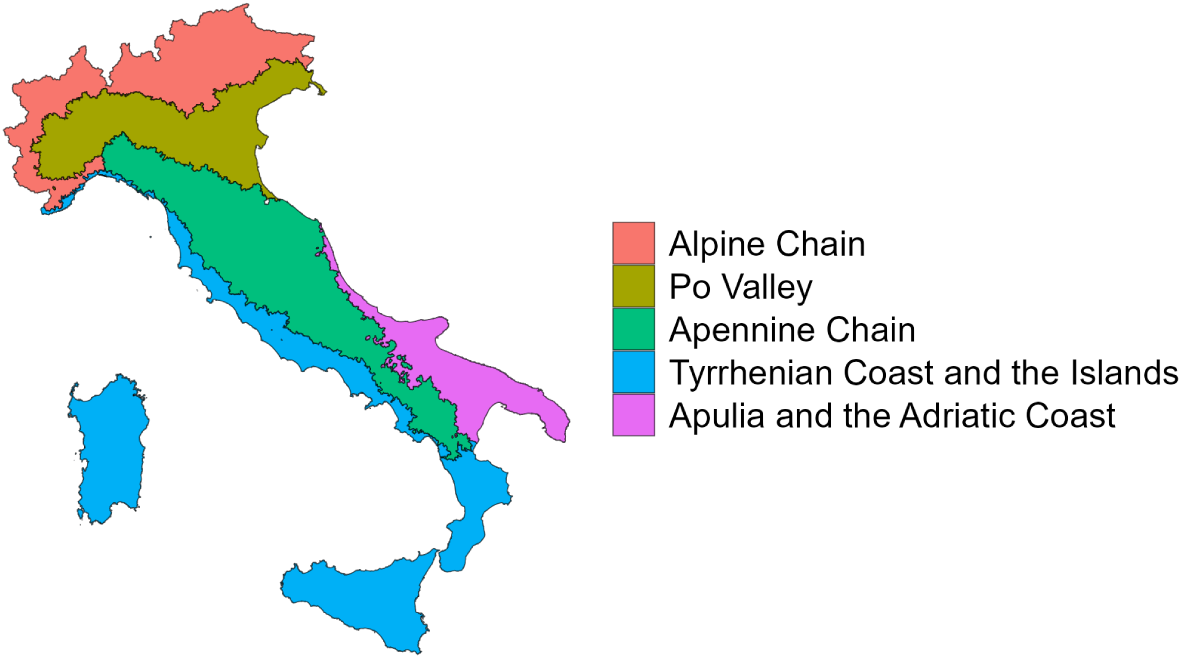
Map of Italy by Ecoregions.

### A.2 Weighted Averages

The spatial units used in the analysis correspond to the municipality level (LAU-2), as of January 1, 2025. In formulas, given {*C*_1_*, …, C*_p_*, …, C*_P_ }, the *P* ERA5-Land grids that fall within Italy’s territory, the temperature for the municipality *s* on day *t* is given by

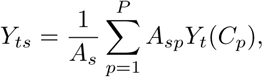

where *A*_s_ is the total surface of the municipality *s*, and *A*_sp_ is the surface of the cell *C*_p_ that falls in the municipality *s*. The total surface of the municipality can also be expressed as 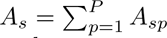, which is the sum of the proportion of the grid cells within its borders.

Missing data for municipality level extractions of exposure covariates were imputed as the mean from the values assigned to their first order neighbours, as defined through the “queen” rule of adjacency.

### A.3 Model Specification

Conception counts *n*_it_, where *i* = 1*, …, I*_eco_ indexes the municipalities belonging to each of the 5 Ecoregions *eco* = 1*, …,* 5, and *t* indexes the LMP week, were modelled through Negative Binomial distribution with a log-link function in a hierarchical Bayesian framework.

The full model can be written as

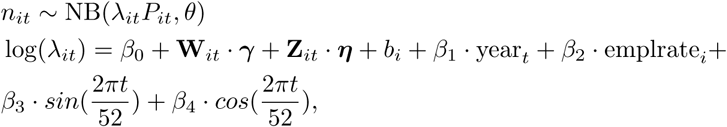

where, *θ* is the overdispersion parameter, and *P*_it_ represents the population of municipality *i* interpolated at the *t*-th week and included as an offset so that *lambda*_it_ can be interpreted as a birth rate from LMP.

The model includes a global intercept *β*_0_, which represents the average birth rate across the country for the entire study period. The term **W**_it_ · ***γ*** represents the non linear effect of temperature across the 3–21 weeks following the last menstrual period and it is modelled using the Distributed Lag Non-Linear model (DLNM) specification [18], so that **W**_it_ are cross-basis functions for the combination of temperature and gestational weeks while ***γ*** are the fixed effects coefficients. A second DLNM component is specified on **Z**_it_·***η*** which captures the effect of NO_2_ across the same period. Differently from the temperature DLNM, here the effect is modelled linearly for each gestational week, but non-linearity is allowed across gestational weeks.

All non-linear relationships were specified using natural cubic splines. For both cross-basis functions, the lead dimension was modelled including an intercept and 3 degrees of freedom over the 3–21 weeks following the LMP. For temperature, knots were placed at the 10^th^ and 90^th^ percentiles of the Ecoregion-specific temperature distribution, for a total of 3 degrees of freedom. The term *β*_1_ captures the linear yearly trend, and *β*_2_ estimates the effect of the municipality-level employment rate. The terms *β*_3_ captures and *β*_4_ capture respectively the sine and cosine Fourier terms with period of 52 weeks.

Lastly we include a spatial and a temporal random effects. The first, *b*_i_, is specified for each municipality and is modelled through a Besag–York–Mollié 2 (BYM2) [15] as a combination of residual structured and unstructured spatial effect, where the structured is defined based on neighbourhoods; the latter, *ω*_t_, captures the temporal seasonal structure, and is defined through a pair of sine-cosine terms, assuming one peak per year.

### A.4 Prior Specification

We now specify the random effects formulations. The BYM2 can be expressed as follows:

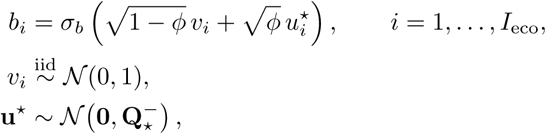

where *u*^★^ is a spatially structured component specified by an intrinsic conditional autoregressive (ICAR) prior, and *v*_i_ is an unstructured latent component with Gaussian prior specification. *ϕ* is the mixing parameter that indicates the proportion of marginal variance explained by either component. Finally, *σ*_b_ is the is the standard deviation of the spatial field.

The RW2 is specified as follows:

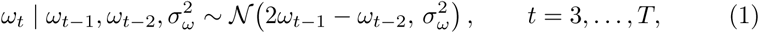

where *σ*^2^ is the innovation variance of the RW2 process, *ω*_t_*_−_*_1_ and *ω*_t_*_−_*_2_ denote the temporal random effects at the two previous weeks. Both the *b*_i_ and *ω*_t_ were specified using Penalised Complexity (PC) priors [15]. Finally, the priors for the fixed effects have a Gaussian prior by default with zero mean and precision equal to 0.001 [30].

### A.5 Ethical approval and data access

The study is about secondary, aggregated data so no additional ethical permission is required. According to the Italian legislation, the approval by an Ethics Committee is only mandatory for clinical trials on medicinal products for human use (according to Regulation EU 2014/536), for clinical trials on medical devices (According to Regulation EU 2017/745), for observational pharmacological studies with drugs (According to Italian Ministerial Decree of November 30, 2021). For other type of studies the approval by an ethics committee, although recommended, is not legally mandatory. Data from the Italian Birth Registry’s “Certificate of Childbirth Assistance” (CeDAP) are available from the Istituto Superiore di Sanità (ISS), but access to them is subject to restrictions; these data were used under license for the purposes of this study and are therefore not publicly accessible. However, the data are available from the authors upon receipt of a justified request and subject to authorization by the ISS. Requests for access to the CeDaP dataset should be sent to.

## B Supplementary Tables

### B.1 Descriptive analysis

#### B.1.1 Attrition table

The attrition table is presented below:

**Supplementary Figure 1:** Attrition table: Sample selection (percentage drop at Each Step)

| Step | Description | Count | Drop (%) | Cum. Drop (%) |
| --- | --- | --- | --- | --- |
| 1 | Initial number of births | 6885869 | 0.000 | 0.000 |
| 2 | LMP 01-01-2013 onwards | 4754787 | 30.949 | 30.949 |
| 3 | Gestational Weeks 22–42 | 4749500 | 0.111 | 31.025 |
| 4 | Maternal age 16–49 years | 4740859 | 0.182 | 31.242 |
| 5 | Mainland Municipality | 4607302 | 2.817 | 34.059 |
| 6 | Single birth | 4457468 | 3.252 | 37.311 |

### B.2 Exposure distributions

**Table 1:** Mean and Standard Deviation of T*mean* by Ecoregion and Italy.

| Ecoregion | Mean | Standard Deviation |
| --- | --- | --- |
| Alpine Chain | 12.96 | 8.37 |
| Po Valley | 18.07 | 8.26 |
| Apennine Chain | 17.48 | 7.97 |
| Tyrrhenian Coast and the Islands | 19.63 | 7.22 |
| Apulia and the Adriatic Coast | 20.10 | 7.70 |
| Italy (Nationwide) | 17.20 | 8.36 |

**Table 2:** Mean and Standard Deviation of N*O*_2_ levels by Ecoregion and Italy.

| Ecoregion | Mean | Standard Deviation |
| --- | --- | --- |
| Alpine Chain | 10.82 | 8.84 |
| Po Valley | 18.48 | 11.65 |
| Apennine Chain | 7.69 | 5.20 |
| Tyrrhenian Coast and the Islands | 6.91 | 6.47 |
| Apulia and the Adriatic Coast | 6.61 | 3.84 |
| Italy | 11.12 | 9.67 |

EC can you make a table like this but for employment rate?

### B.3 Descriptive statistics

**Table 3:** Fixed effects of Employment rate (20–64) by Ecoregion, RR and 95% Credible Interval.

| Ecoregion | RR | 95% CrI |
| --- | --- | --- |
| Po Valley | 0.0106 | (0.0026 - 0.0185) |
| Alpine Chain | 0.0365 | (0.0244 - 0.0485) |
| Apennine Chain | 0.0573 | (0.0411 - 0.0737) |
| Tyrrhenian Coast and the Islands | -0.0441 | (-0.0529 - -0.0352) |
| Apulia and the Adriatic Coast | 0.0046 | (-0.012 - 0.0211) |

**Table 4:** Fixed effects of year by Ecoregion, RR and 95% Credible Interval.

| Ecoregion | RR | 95% CrI |
| --- | --- | --- |
| Po Valley | -0.0151 | (-0.0159 - -0.0143) |
| Alpine Chain | -0.0139 | (-0.0152 - -0.0127) |
| Apennine Chain | -0.0151 | (-0.0161 - -0.0141) |
| Tyrrhenian Coast and the Islands | -0.0100 | (-0.0106 - -0.0093) |
| Apulia and the Adriatic Coast | -0.0124 | (-0.0137 - -0.0112) |

**Table 5:** Percentile–based knot positions of the mean temperature distribution.

| model | 10% | 90% |
| --- | --- | --- |
| Po Valley | 3.41 | 24.10 |
| Alpine Chain | -1.41 | 19.72 |
| Apennine Chain | 4.08 | 22.97 |
| Tyrrhenian Coast and the Islands | 7.76 | 24.78 |
| Apulia and the Adriatic Coast | 7.35 | 25.76 |

#### B.3.1 Birth Rates

### B.4 Model Output

**Table 6:**
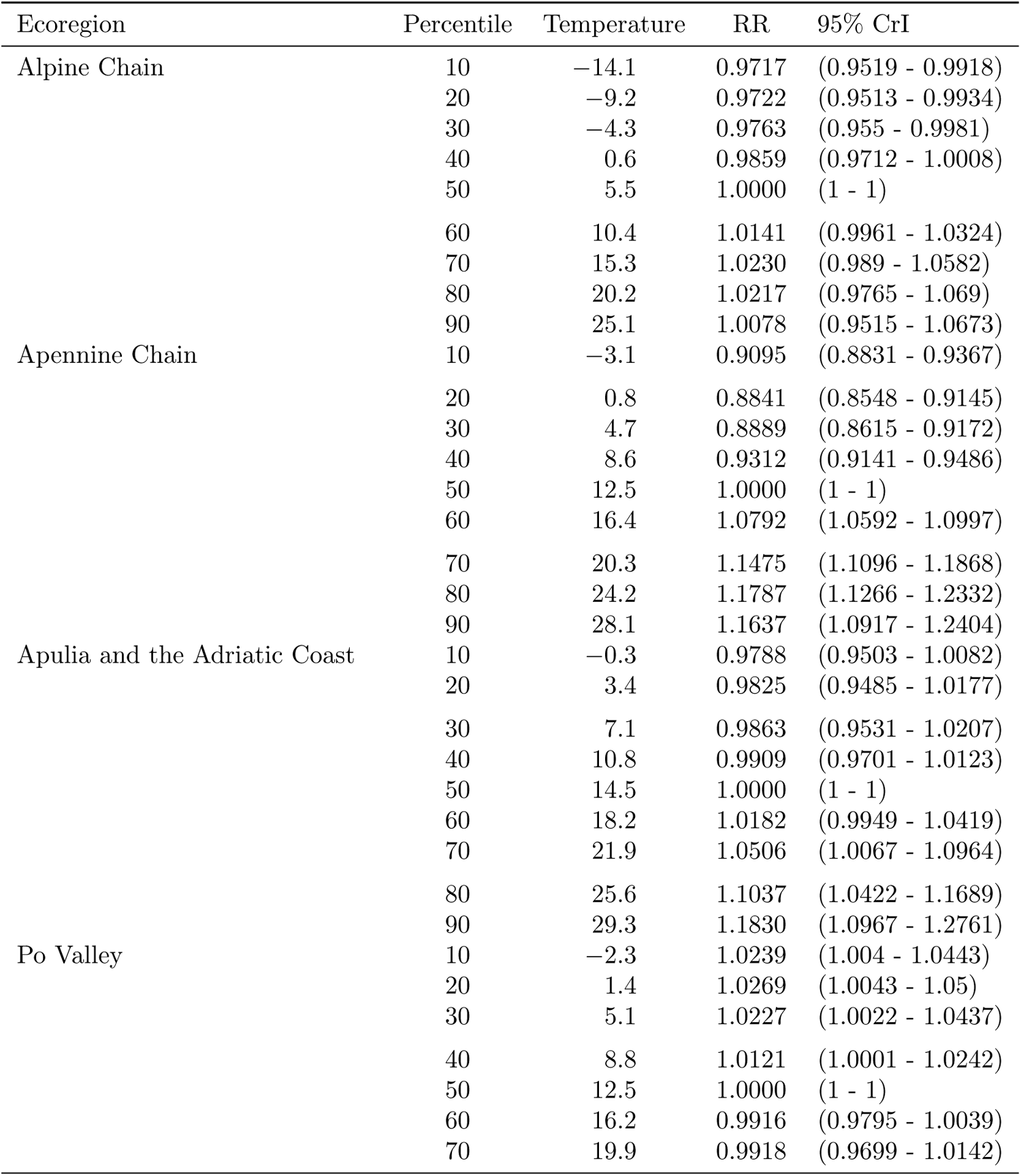
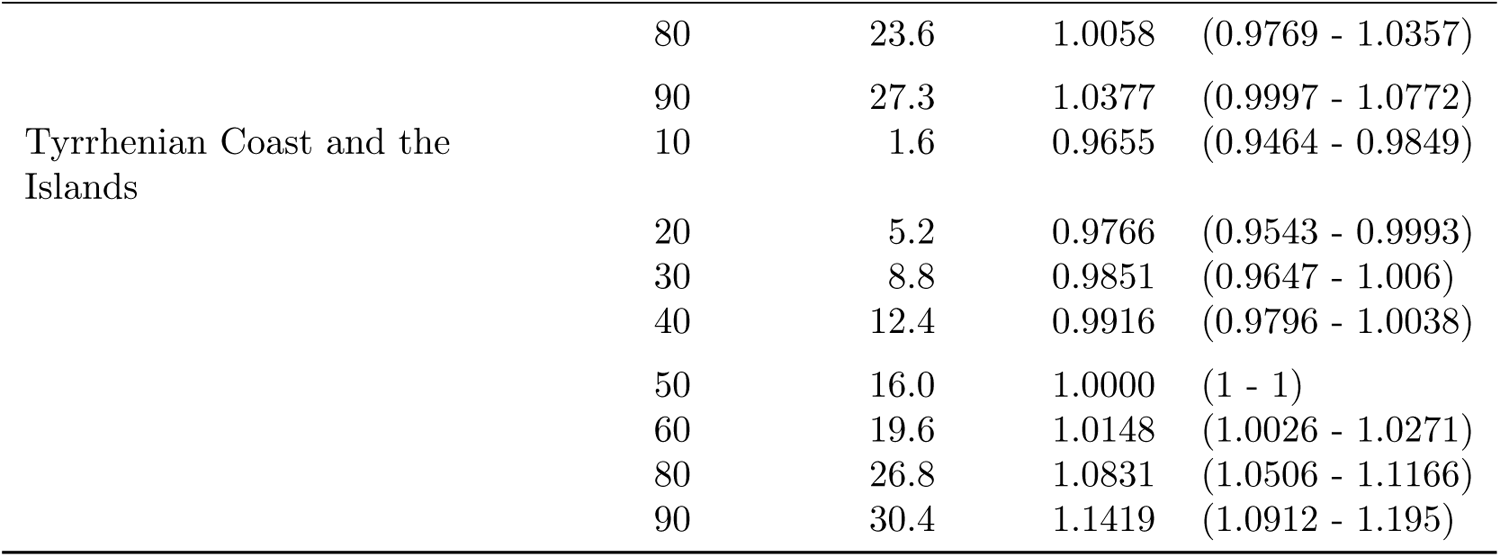
Birth rates in response to temperature exposure throughout GWs 3–21, by decile of mean temperature.

**Table 7:**
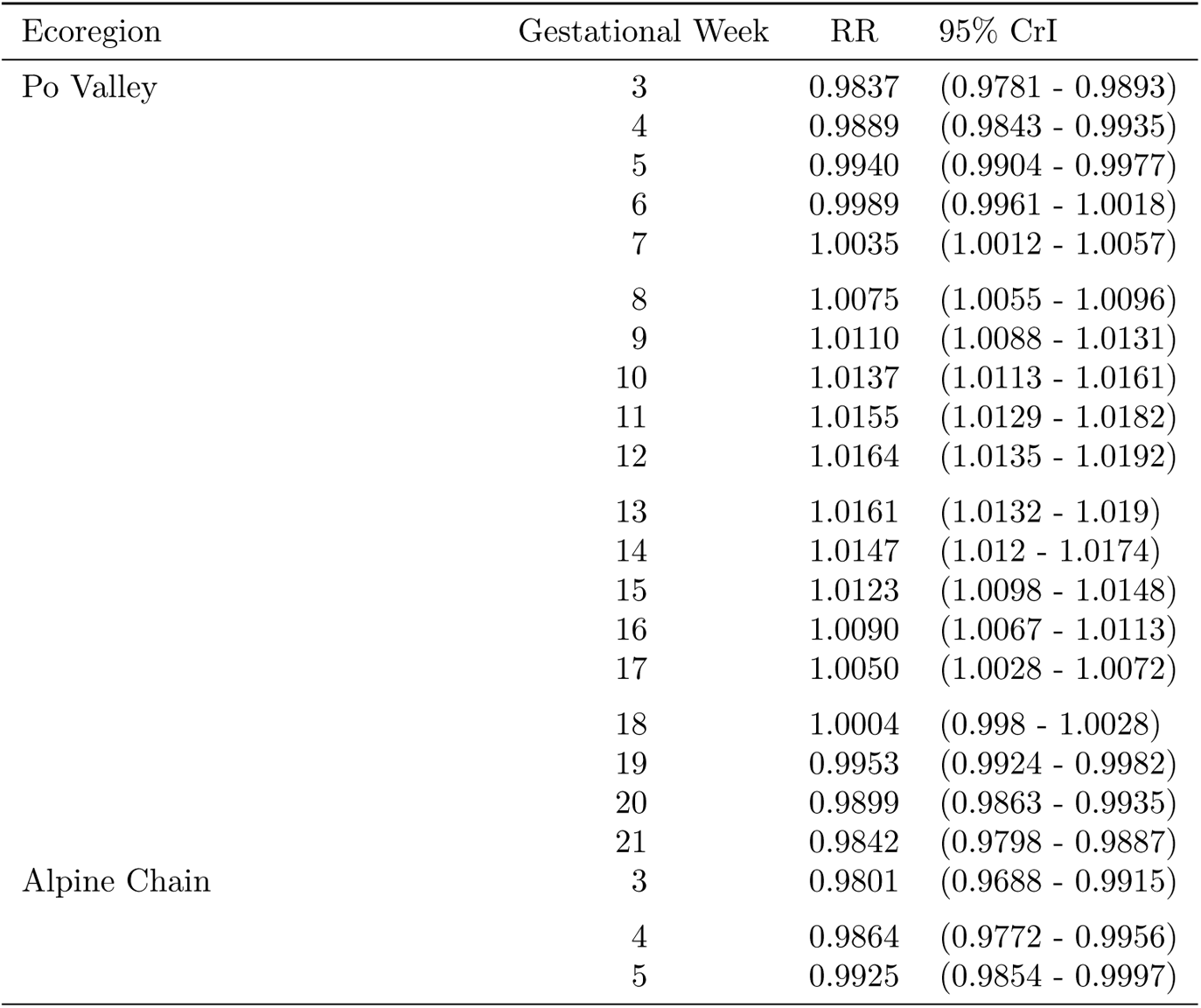
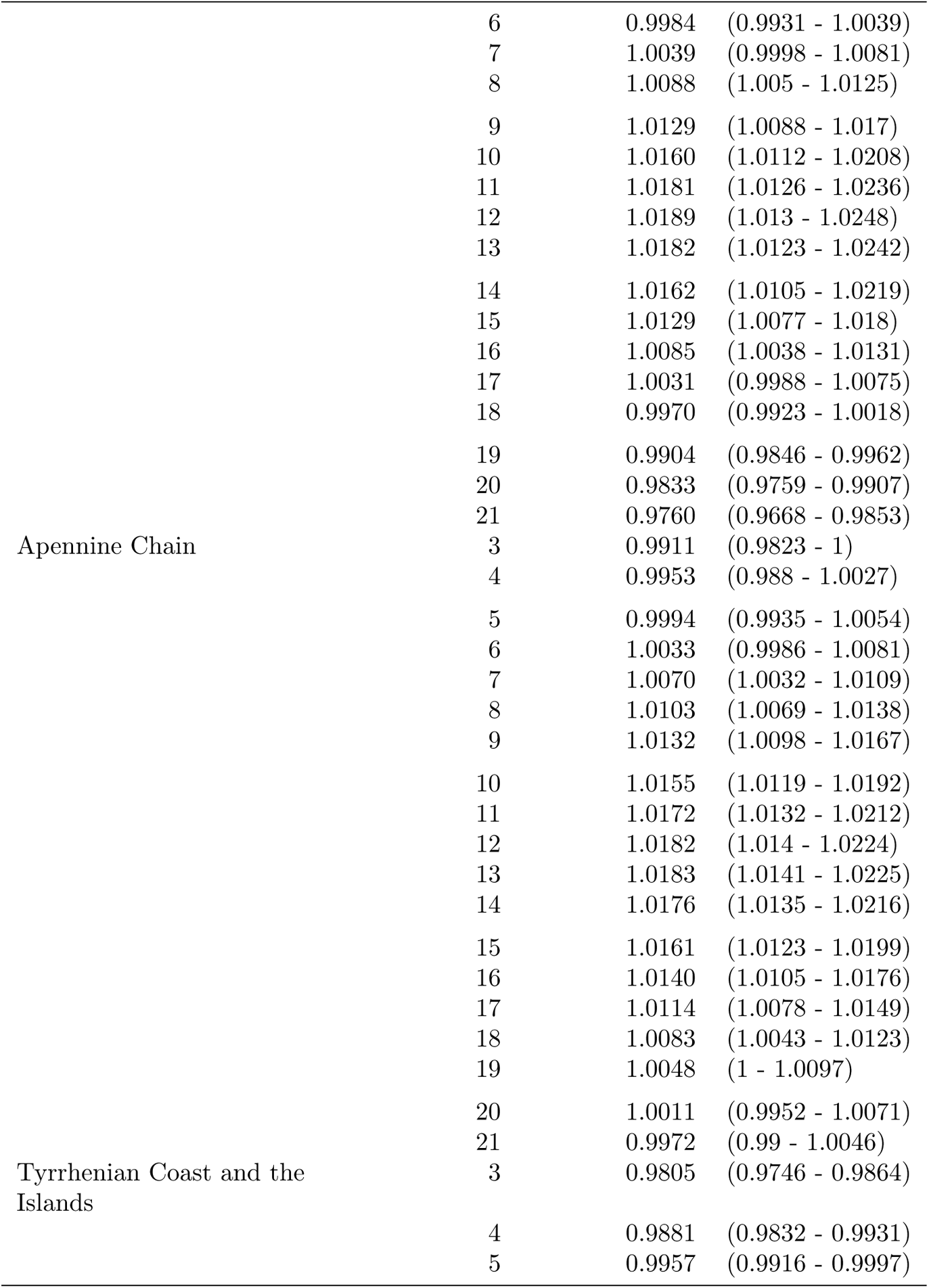
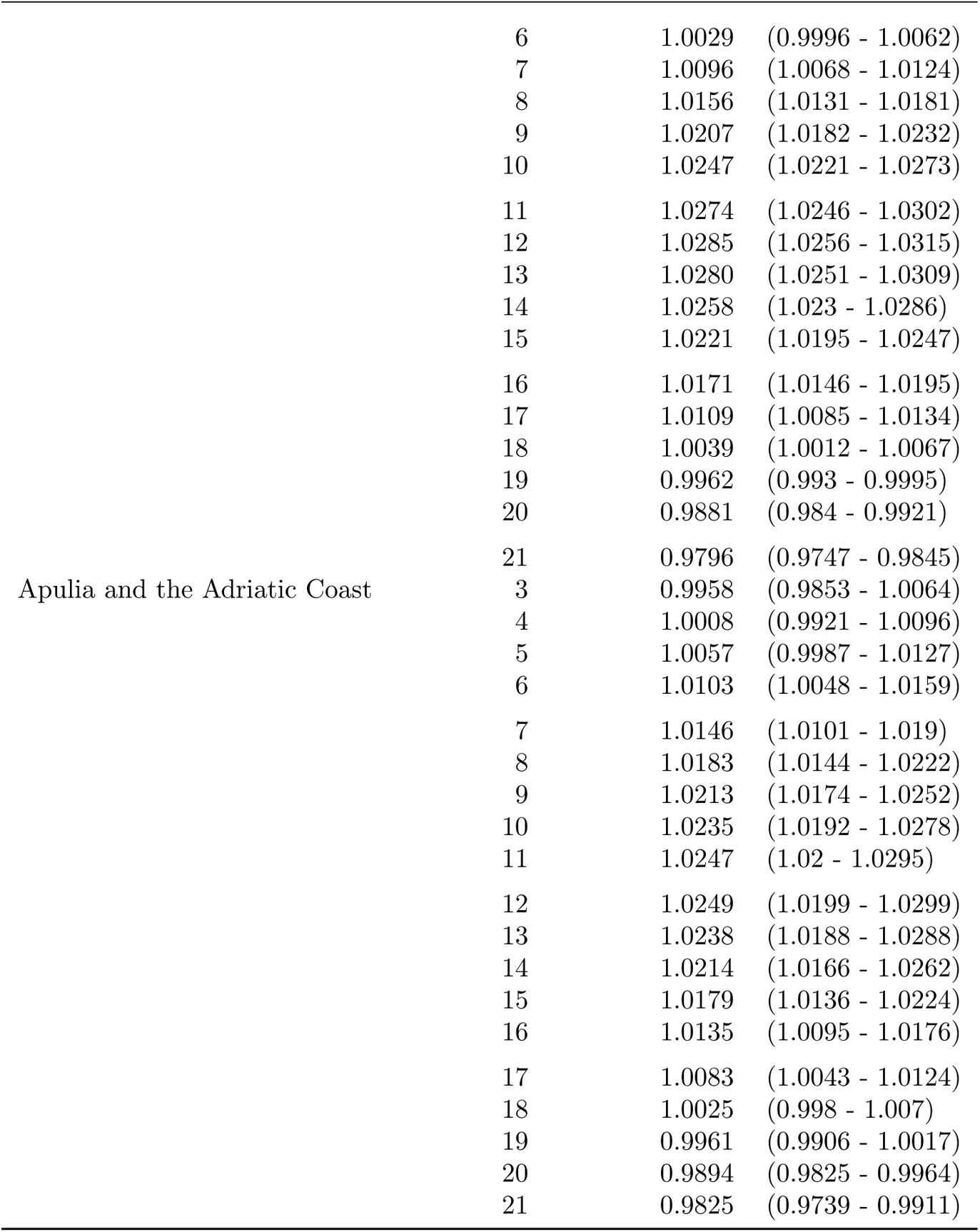
Effect of heat on birth rates by gestational week, defined as relative risk of birth at the 90^th^ percentile relative to the median.

**Table 8:**
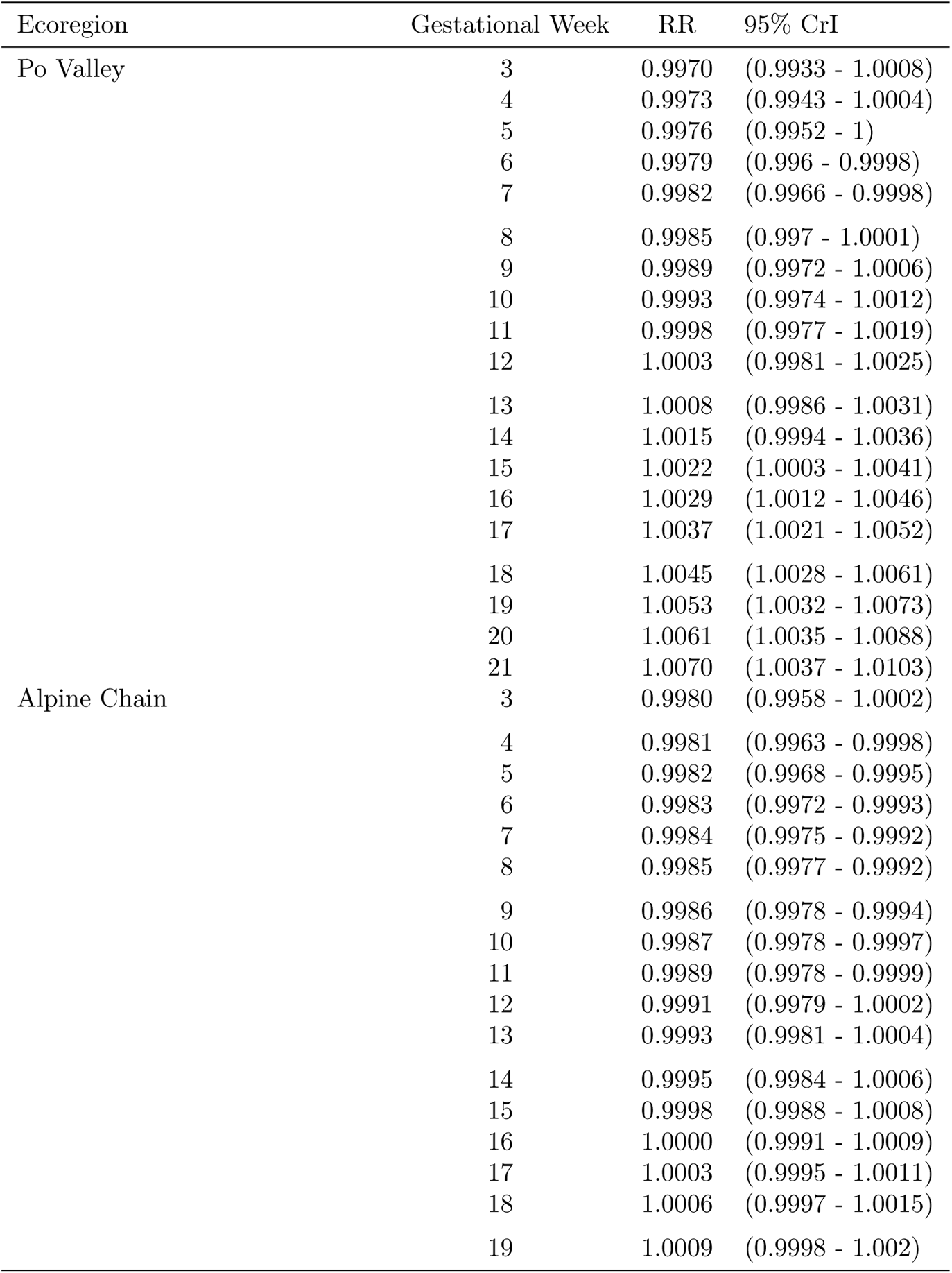
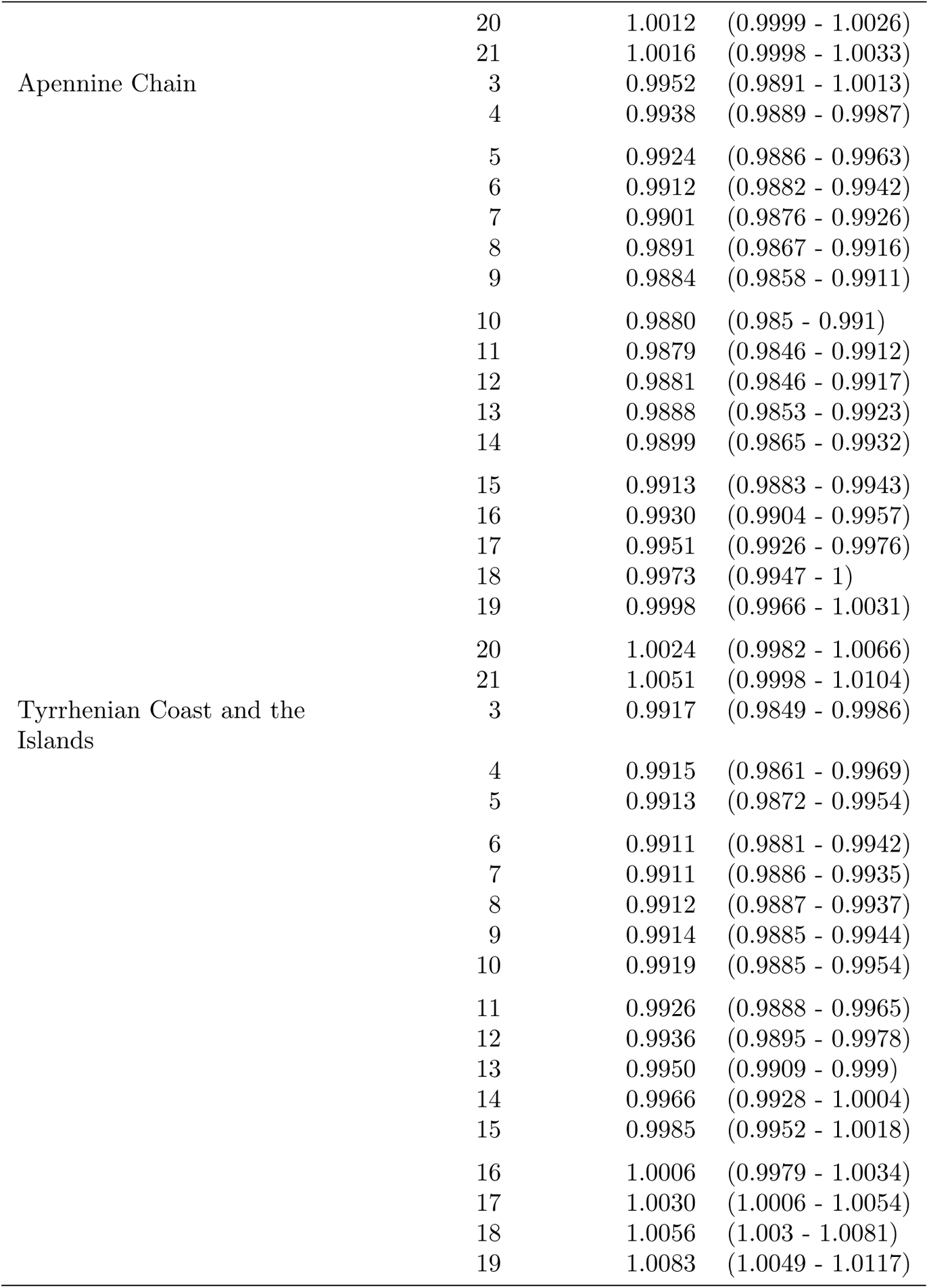
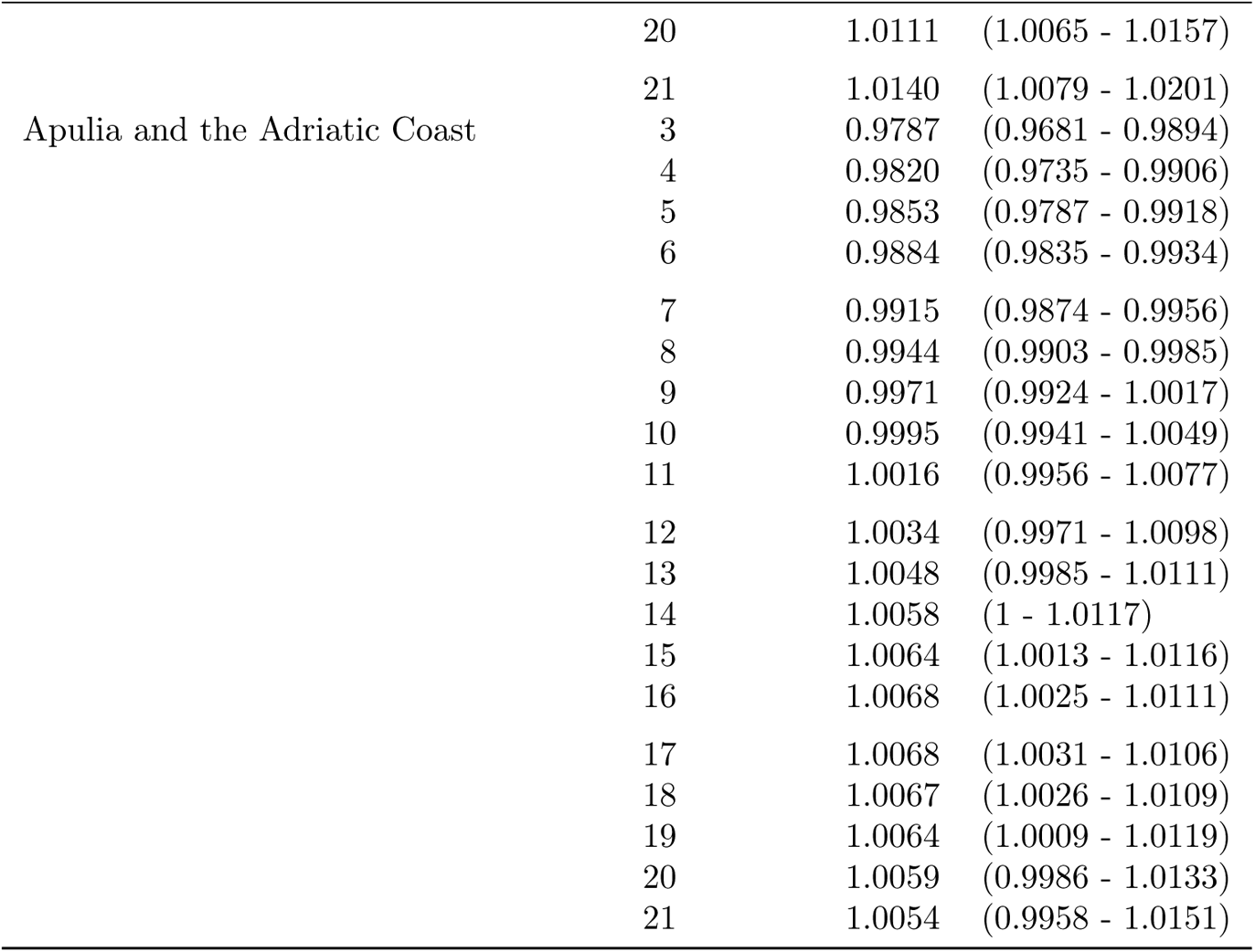
Effect of cold on birth rates by gestational week, defined as relative risk of birth at the 10^th^ percentile relative to the median.

**Table 9:**
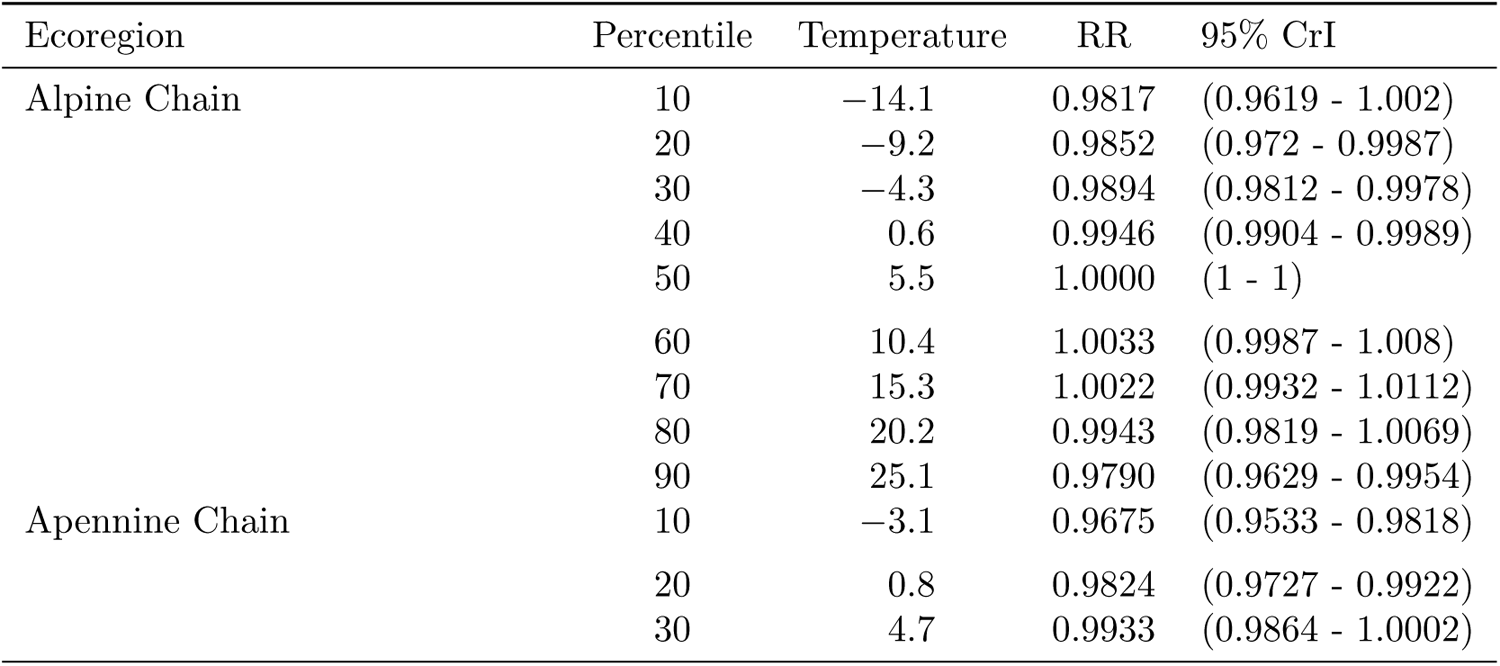
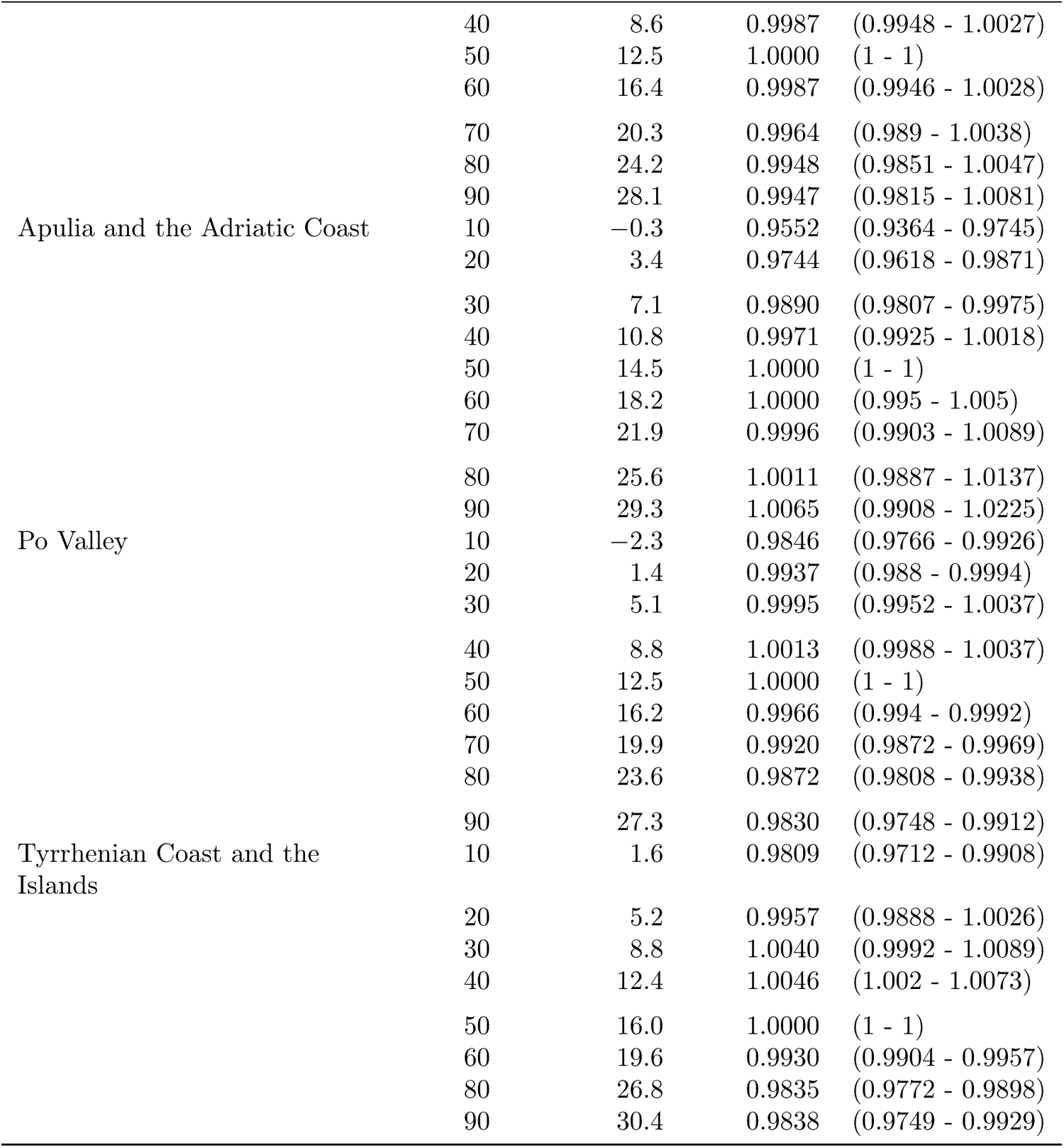
Birth rates in response to temperature exposure throughout GWs 3–4, by decile of mean temperature.

**Table 10:**
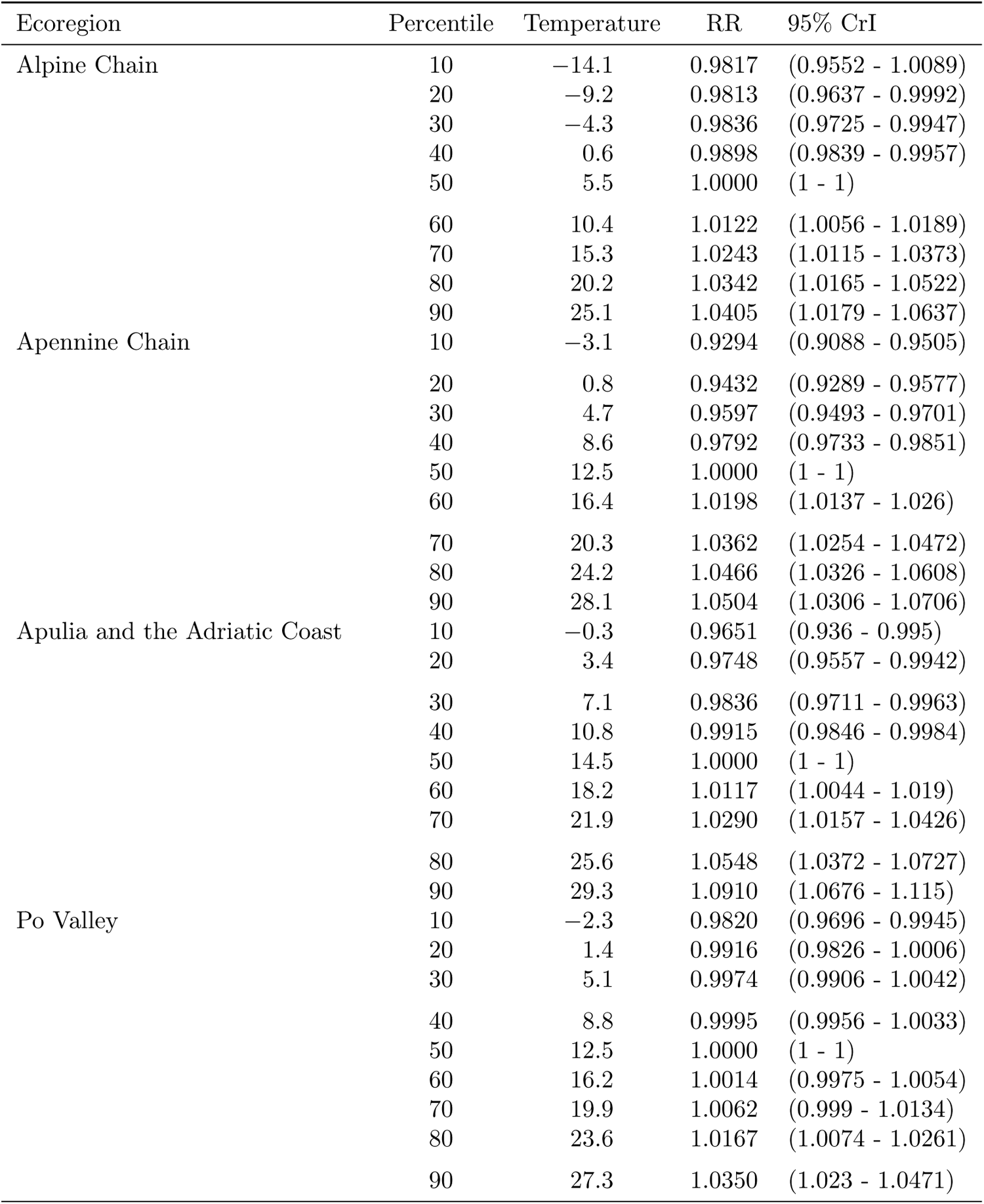
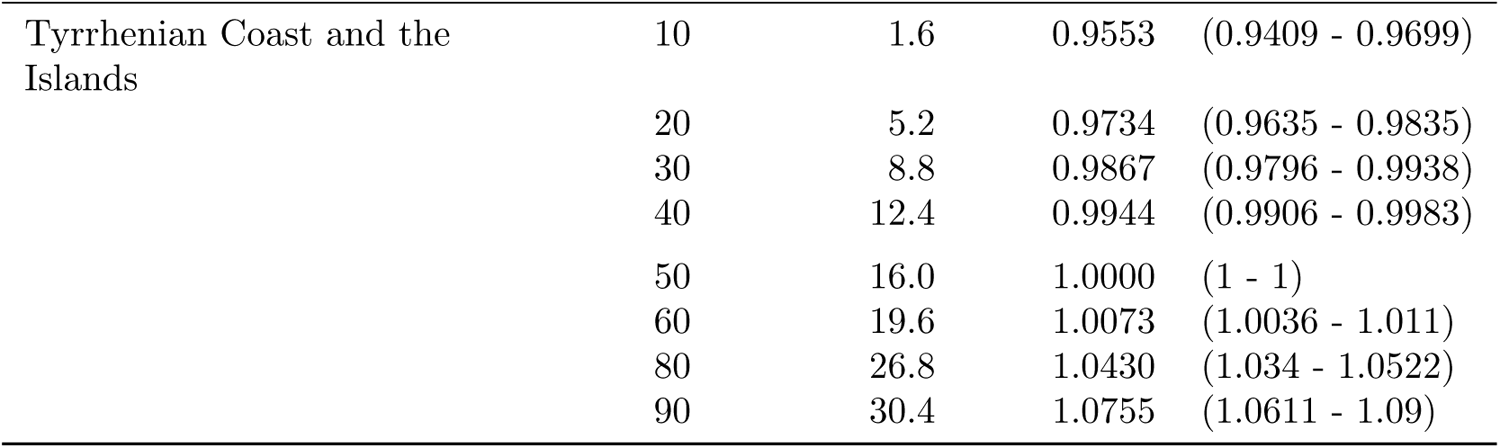
Birth rates in response to temperature exposure throughout GWs 5–9, by decile of mean temperature.

| Ecoregion | Percentile | Temperature | RR | 95% CrI |
| --- | --- | --- | --- | --- |
| Alpine Chain | 10 | −14.1 | 0.9817 | (0.9552 - 1.0089) |
|  | 20 | −9.2 | 0.9813 | (0.9637 - 0.9992) |
|  | 30 | −4.3 | 0.9836 | (0.9725 - 0.9947) |
|  | 40 | 0.6 | 0.9898 | (0.9839 - 0.9957) |
|  | 50 | 5.5 | 1.0000 | (1 - 1) |
|  | 60 | 10.4 | 1.0122 | (1.0056 - 1.0189) |
|  | 70 | 15.3 | 1.0243 | (1.0115 - 1.0373) |
|  | 80 | 20.2 | 1.0342 | (1.0165 - 1.0522) |
|  | 90 | 25.1 | 1.0405 | (1.0179 - 1.0637) |
| Apennine Chain | 10 | −3.1 | 0.9294 | (0.9088 - 0.9505) |
|  | 20 | 0.8 | 0.9432 | (0.9289 - 0.9577) |
|  | 30 | 4.7 | 0.9597 | (0.9493 - 0.9701) |
|  | 40 | 8.6 | 0.9792 | (0.9733 - 0.9851) |
|  | 50 | 12.5 | 1.0000 | (1 - 1) |
|  | 60 | 16.4 | 1.0198 | (1.0137 - 1.026) |
|  | 70 | 20.3 | 1.0362 | (1.0254 - 1.0472) |
|  | 80 | 24.2 | 1.0466 | (1.0326 - 1.0608) |
|  | 90 | 28.1 | 1.0504 | (1.0306 - 1.0706) |
| Apulia and the Adriatic Coast | 10 | −0.3 | 0.9651 | (0.936 - 0.995) |
|  | 20 | 3.4 | 0.9748 | (0.9557 - 0.9942) |
|  | 30 | 7.1 | 0.9836 | (0.9711 - 0.9963) |
|  | 40 | 10.8 | 0.9915 | (0.9846 - 0.9984) |
|  | 50 | 14.5 | 1.0000 | (1 - 1) |
|  | 60 | 18.2 | 1.0117 | (1.0044 - 1.019) |
|  | 70 | 21.9 | 1.0290 | (1.0157 - 1.0426) |
|  | 80 | 25.6 | 1.0548 | (1.0372 - 1.0727) |
|  | 90 | 29.3 | 1.0910 | (1.0676 - 1.115) |
| Po Valley | 10 | −2.3 | 0.9820 | (0.9696 - 0.9945) |
|  | 20 | 1.4 | 0.9916 | (0.9826 - 1.0006) |
|  | 30 | 5.1 | 0.9974 | (0.9906 - 1.0042) |
|  | 40 | 8.8 | 0.9995 | (0.9956 - 1.0033) |
|  | 50 | 12.5 | 1.0000 | (1 - 1) |
|  | 60 | 16.2 | 1.0014 | (0.9975 - 1.0054) |
|  | 70 | 19.9 | 1.0062 | (0.999 - 1.0134) |
|  | 80 | 23.6 | 1.0167 | (1.0074 - 1.0261) |
|  | 90 | 27.3 | 1.0350 | (1.023 - 1.0471) |
| Tyrrhenian Coast and the Islands | 10 | 1.6 | 0.9553 | (0.9409 - 0.9699) |
|  | 20 | 5.2 | 0.9734 | (0.9635 - 0.9835) |
|  | 30 | 8.8 | 0.9867 | (0.9796 - 0.9938) |
|  | 40 | 12.4 | 0.9944 | (0.9906 - 0.9983) |
|  | 50 | 16.0 | 1.0000 | (1 - 1) |
|  | 60 | 19.6 | 1.0073 | (1.0036 - 1.011) |
|  | 80 | 26.8 | 1.0430 | (1.034 - 1.0522) |
|  | 90 | 30.4 | 1.0755 | (1.0611 - 1.09) |

**Table 11:**
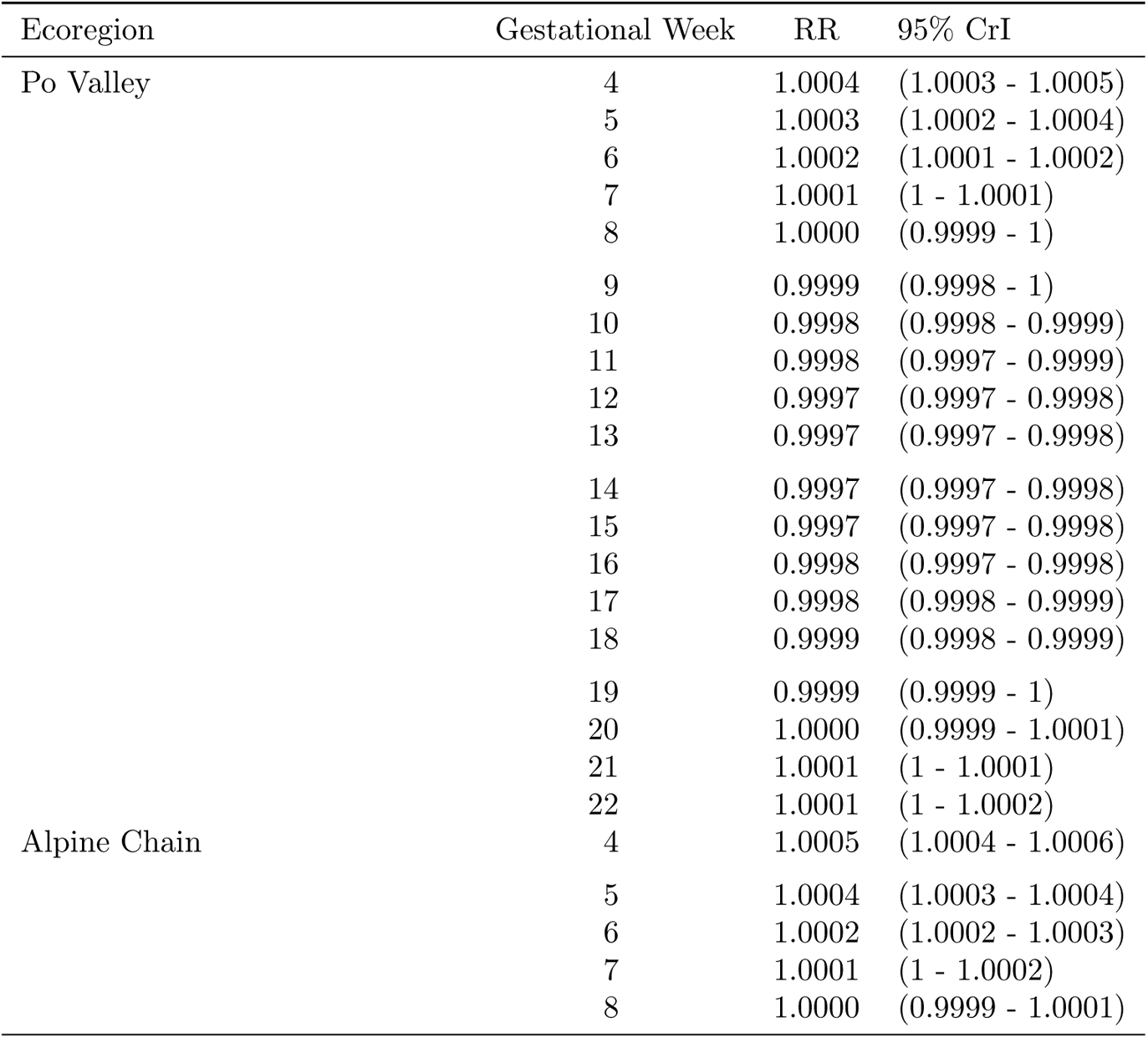
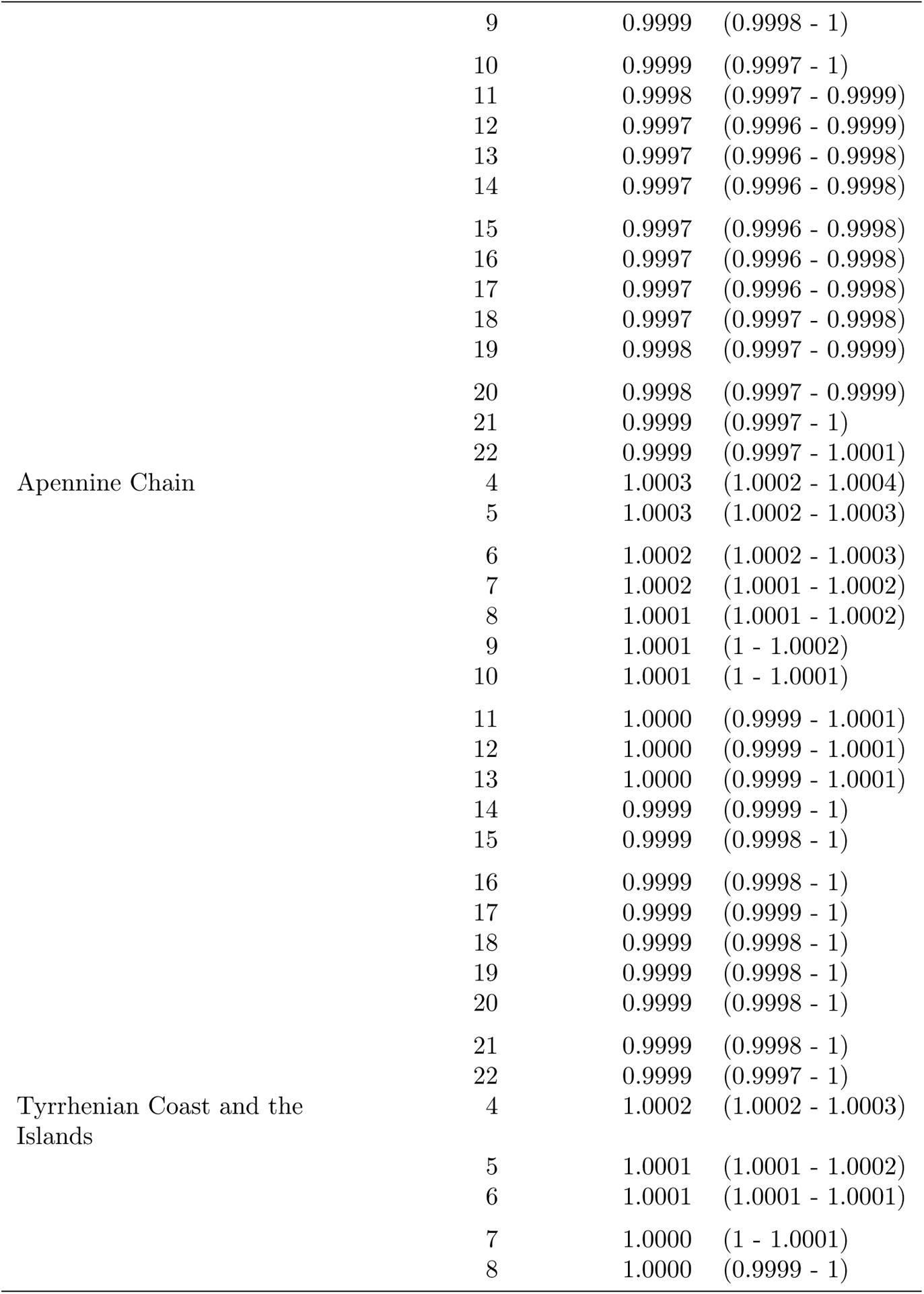
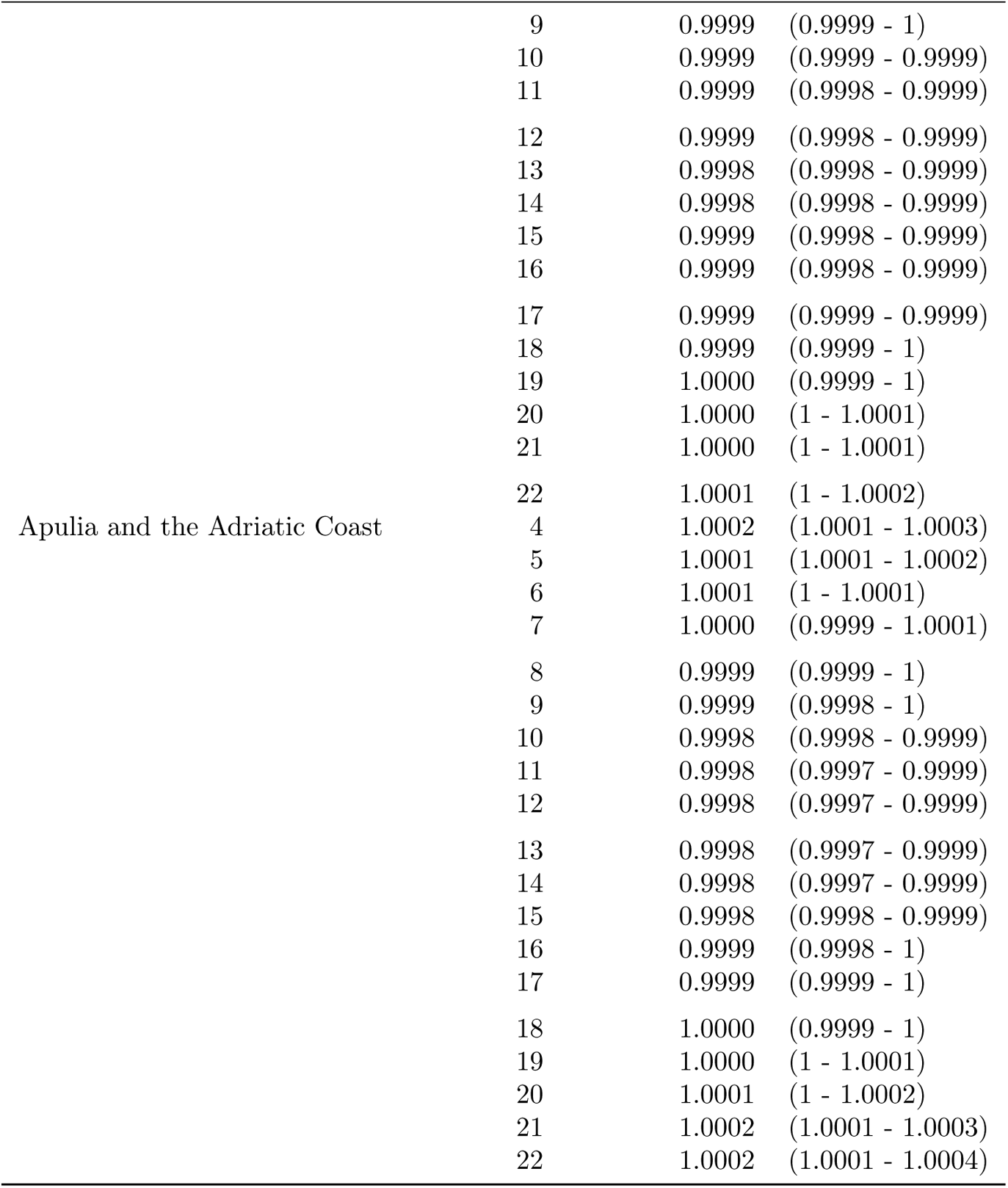
Effect of N*O*_2_ on birth rates by gestational week, defined as relative risk of birth above the 80^th^ percentile.

## C Supplementary Figures

### C.1 Descriptive analysis

**Figure C.1:**
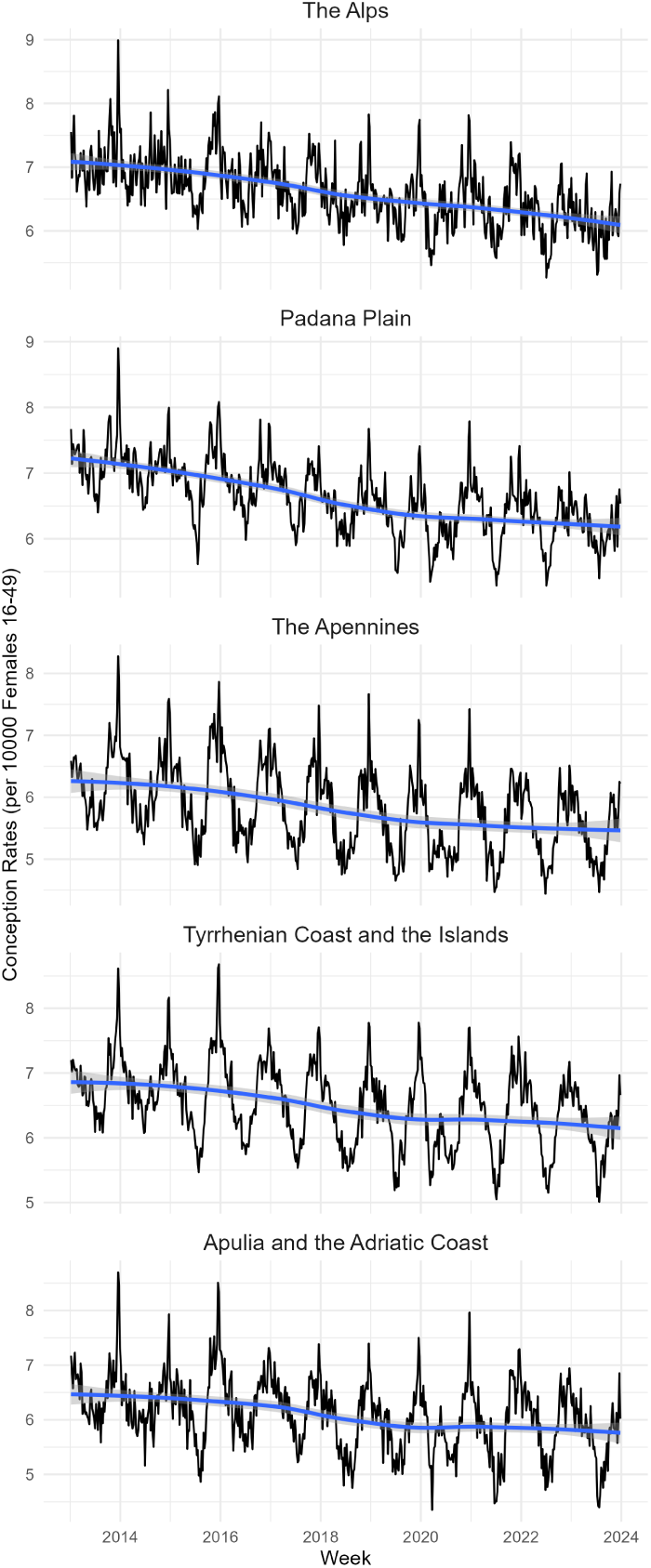
Time series of birth rates from LMP by Ecoregion.

**Figure C.2:**
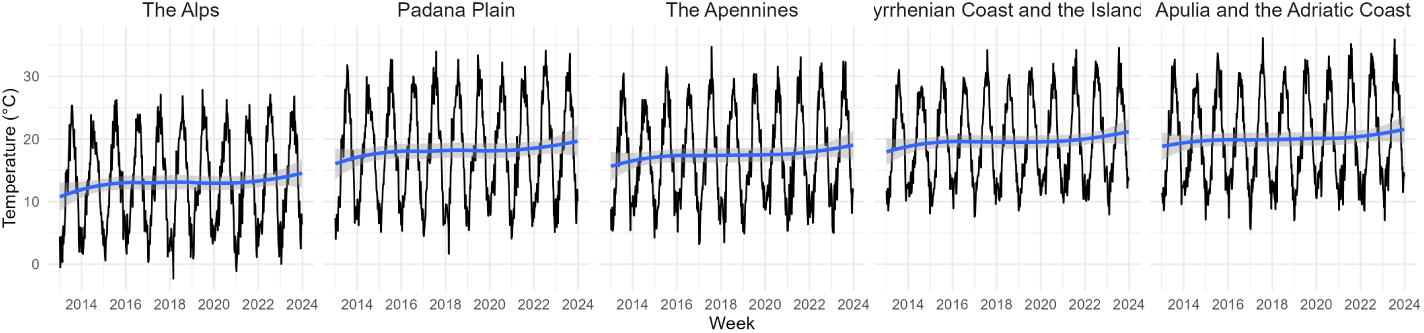
Time series of Average Maximum Weekly Temperature by Ecoregion.

**Figure C.3:**
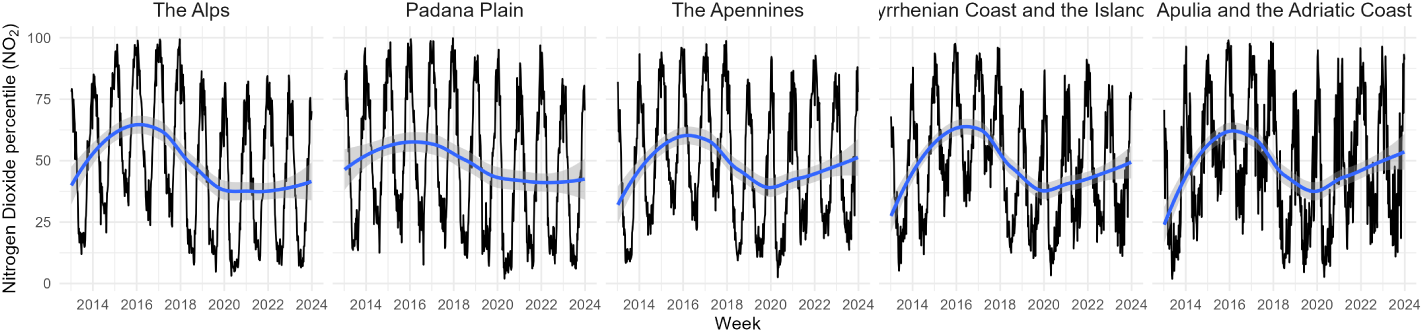
Weekly mean NO_2_ concentrations (*µg/m*^3^) plotted as a time series by Ecoregion.

**Figure C.4:**
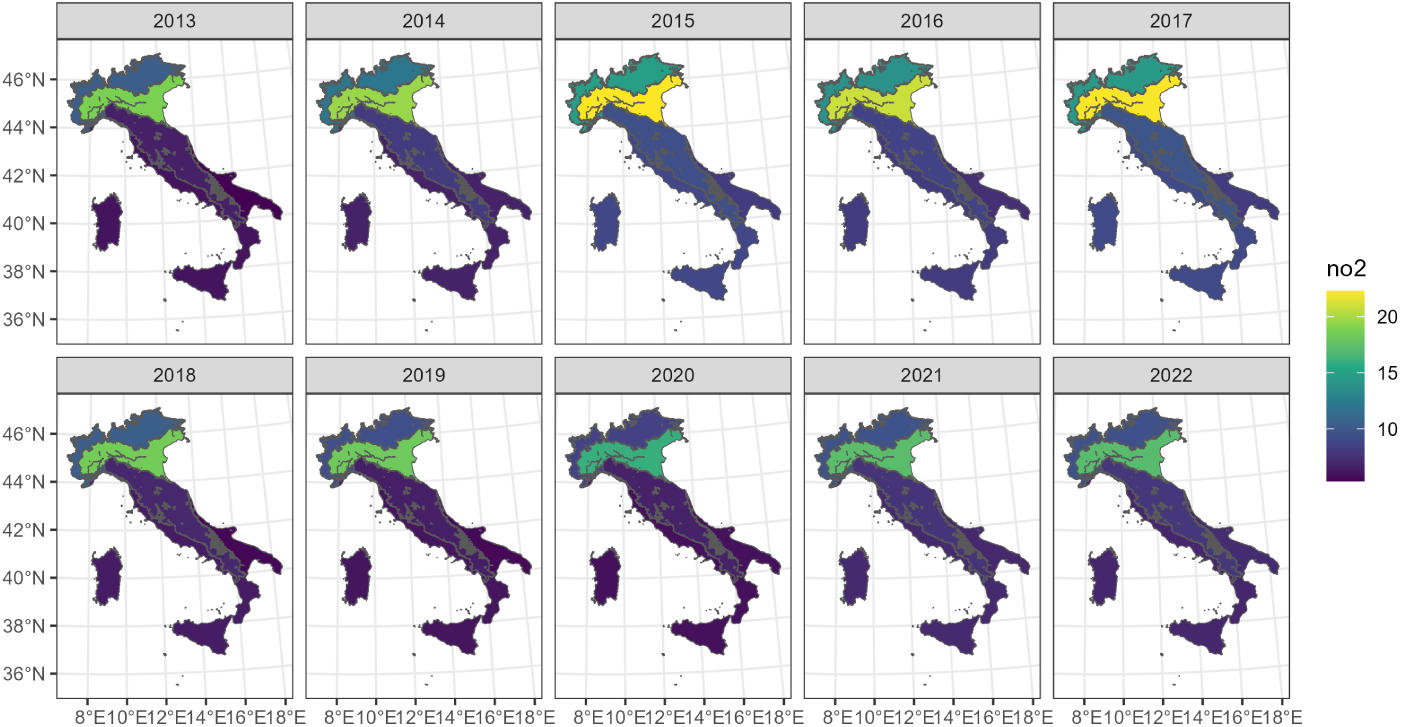
Map of mean NO_2_ concentrations (*µg/m*^3^) by Ecoregion.

### C.2 Model Output

**Figure C.5:**
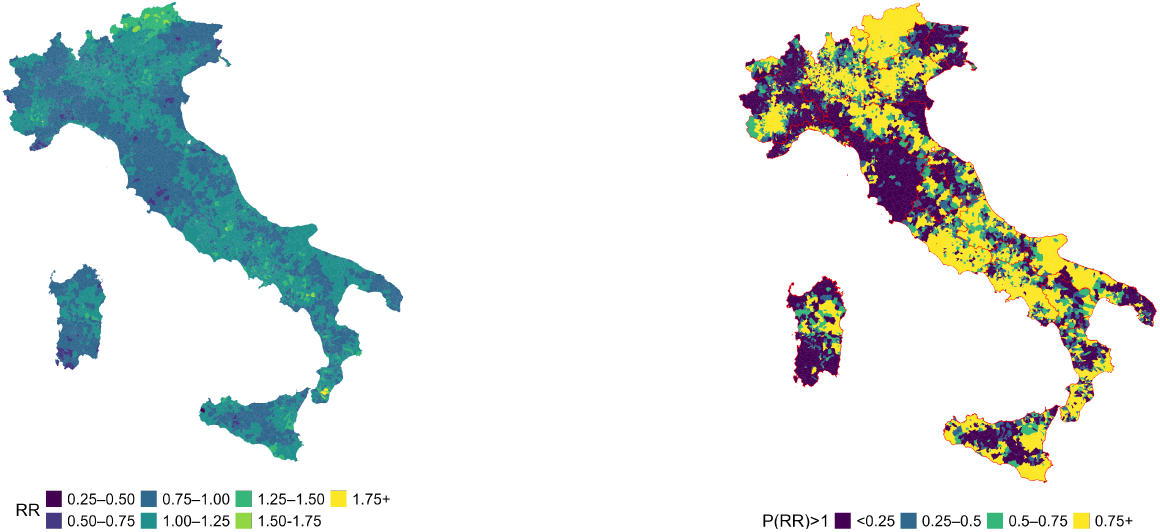
Municipality-level birth rate relative to the national average (left); and posterior probability that relative risk exceeds 1 (right)

**Figure C.6:**
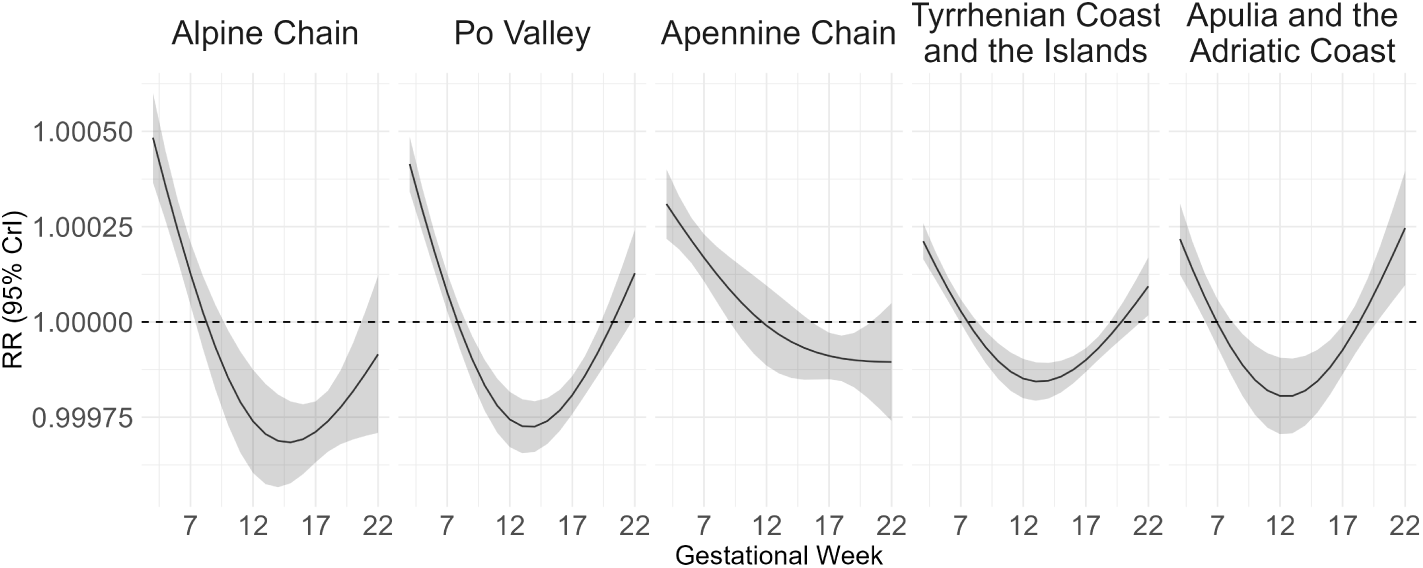
Gestational Week specific association between viable pregnancy from LMP and NO_2_ exposure above the Ecoregion specific 80^th^ percentile.

## D Sensitivity analysis

### D.1 Methods

We tested outcome sensitivity to key modelling assumptions. First, we ran the model excluding preterm births (gestational age *<* 37 weeks). This was to test the possibility that early–gestation temperature exposures may result in variations in the pre–term to full–term birth ratios, which could in turn change the association between temperature and birth rates. We then tested an alternative approach to modelling seasonality, replacing the Fourier sine-cosine term with a second-order Random Walk. We had to ensure that the approach for modelling seasonality was adequate and would not change the exposure-outcome association, because one of the main assumption of the model is that temperature exposure during early gestation may explain fluctuations in births that would not be otherwise explained by seasonal patterns. We tested whether the operationalisation of temperature exposure – the weekly average of daily maxima and daily minima – would yield substantially different results compared to the primary specification. Finally, we tested alternative spline specifications for the temperature–response curve, using natural cubic splines with 4 degrees of freedom, with knots placed at the 10th, 50th and 90th percentiles, respectively.

### D.2 Results

Below are the plots in which the association between birth rates and overall average temperature during GWs 3-22 of the main model are compared to the results obtained from the sensitivity analyses, as described in Supplementary Section D.1.

Supplementary Figure D.1 displays the shape of the association between using mean, maximum and minimum daily temperatures in the weekly average aggregation. The shape of the association remains constant. The most prominent differences can be observed in the Po Valley, where minimum temperature results in estimates closest to null, and the most constant association is to be found in the Alpine Chain region. In the Apennine, Thyrrhenian and Adriatic Regions, minimum temperature gives the highest increase of birth rates for warmer temperatures. When comparing different methods of modelling seasonality, the differences in overall association are minimal, and almost perfectly overlapping curves are produced for the Thyrrhenian Coast Ecoregion. The Apennine Chain gets a higher peak of birth rates in response to warmer temperatures for the sine-cosine model, but the RR fall within each other’s Credible Intervals D.3. Similarly, overlapping curves are found when comparing the models which included births at GWs 22–42 and full term only D.4. The models with 4 degrees of freedom resulted in consistent curves across all Ecoregions but the Apennine Chain, where adding the extra degree of freedom prevented the curve from dipping at the highest range of the temperature domain D.2.

**Figure D.1:**
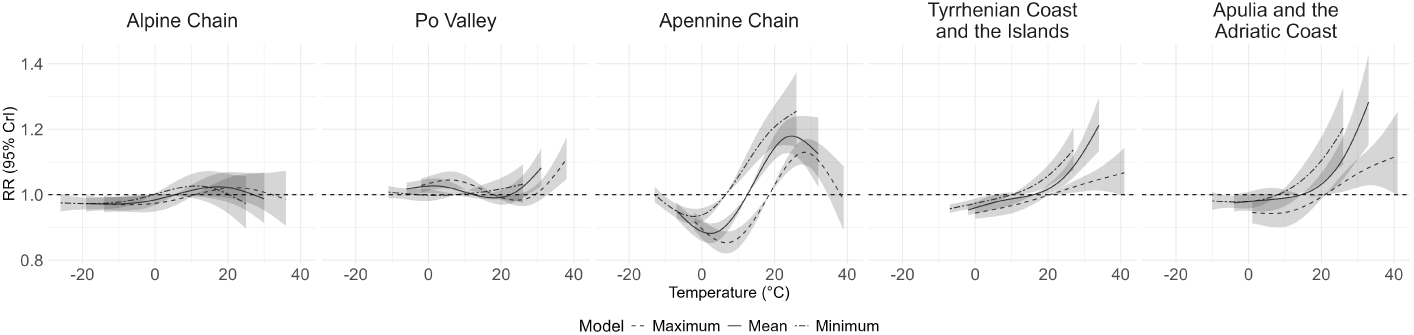
Comparison of sensitivity to different temperature operationalisation: weekly mean of daily mean, daily minimum, daily maximum.

**Figure D.2:**
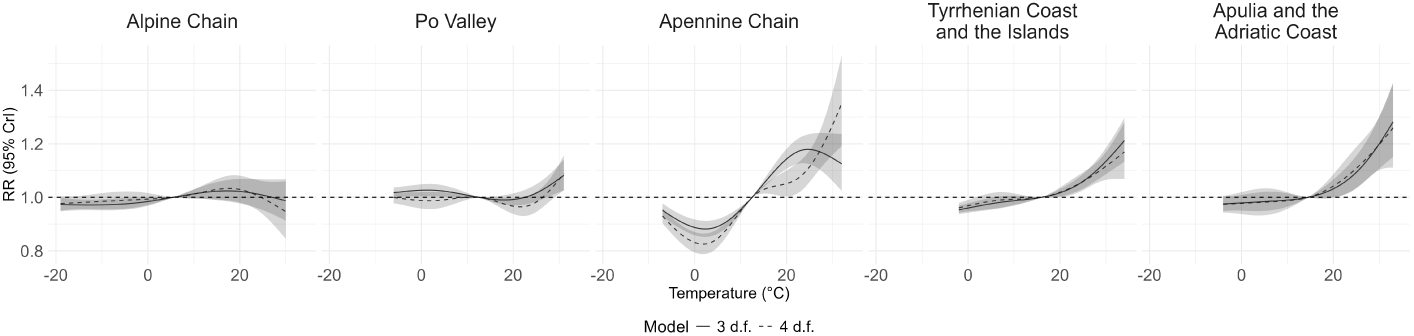
Comparison of sensitivity to different knot positions of the mean temperature distribution: 3 d.f. (10th, 90th percentiles) 4 d.f. (10th, 50th, 90th percentiles).

**Figure D.3:**
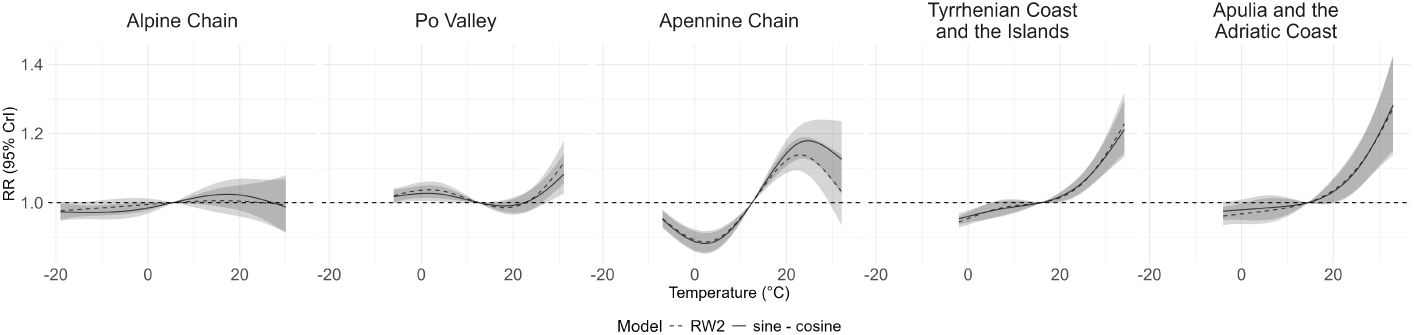
Comparison of sensitivity to different modelling approaches for seasonality: one pair of sine-cosine terms, and random walk of second order (RW2).

**Figure D.4:**
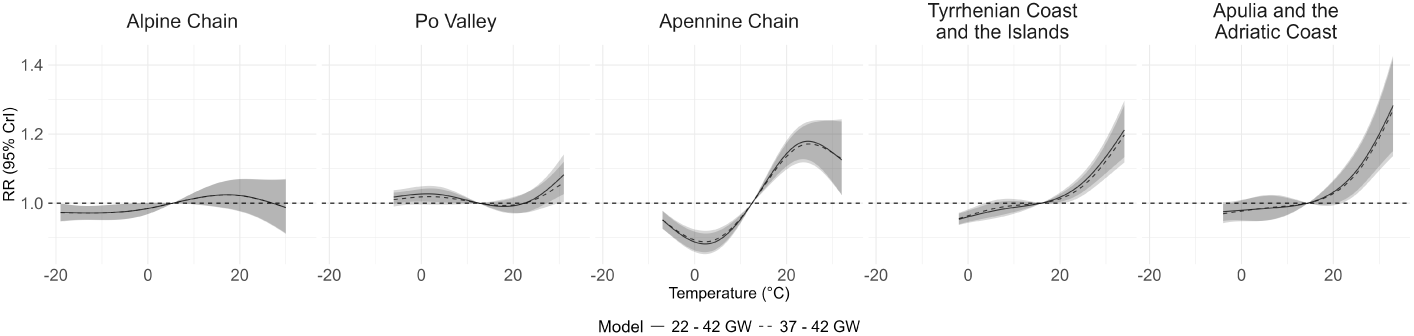
Comparison of sensitivity to different sample selection: GWs 22–42 and full term only, GWs 37–42.

## Notes

### Competing Interest Statement

The authors have declared no competing interest.

### Author Declarations

Ethics committee of Unita di Bioetica, Istituto Superiore di Sanita (ISS) waived ethical approval of this work, based on the following: According to the Italian legislation, the approval by an Ethics Committee is only mandatory for clinical trials on medicinal products for human use (according to Regulation EU 2014/536), for clinical trials on medical devices (According to Regulation EU 2017/745), for observational pharmacological studies with drugs (According to Italian Ministerial Decree of November 30, 2021)

## References

1. Samuels L, Nakstad B, Roos N, Bonell A, Chersich M, Havenith G, Luchters S, Day LT, Hirst JE, Singh T, Elliott-Sale K, Hetem R, Part C, Sawry S, Le Roux J, and Kovats S. Physiological mechanisms of the impact of heat during pregnancy and the clinical implications: review of the evidence from an expert group meeting. International Journal of Biometeorology 2022; 66:1505–13. doi: 10.1007/s00484-022-02301-6. Available from: https://pmc.ncbi.nlm.nih.gov/articles/PMC9300488/ [Accessed on: 2026 Apr 12]

2. Donovan MF and Cascella M. Embryology, Weeks 6-8. eng. StatPearls. Treasure Island (FL): StatPearls Publishing, 2025. Available from: http://www.ncbi.nlm.nih.gov/books/NBK563181/ [Accessed on: 2025 Oct 21]

3. Iachino A, Boldrini R, Basili F, Bergamaschi A, Campo G, Di Cesare M, Moroni R, Rizzuto E, Romanelli M, and Trevisani V. Analisi dell’evento nascita. Anno 2024. Pubblicazione statistica. Disponibile nella sezione “Pubblicazioni principali del Sistema statistico sanitario”. Roma: Ministero della Salute – Ufficio di Statistica, 2024. Available from: https://www.salute.gov.it/statistiche

4. Doing a pregnancy test. en. 2020 Dec. Available from: https://www.nhs.uk/pregnancy/trying-for-a-baby/doing-a-pregnancy-test/ [Accessed on: 2025 Oct 31]

5. Branum AM and Ahrens KA. Trends in Timing of Pregnancy Awareness Among US Women. eng. Maternal and Child Health Journal 2017 Apr; 21:715–26. doi: 10.1007/s10995-016-2155-1

6. ISTAT. Comuni secondo le Ecoregioni d’Italia. Published on 18 dicembre 2018; updated on 12 ottobre 2023; acccessed on: 12 January 2026. 2023. Available from: https://www.istat.it/statistica-sperimentale/classificazione-dei-comuni-secondo-le-ecoregioni-ditalia/

7. IPCC - Intergovernmental Panel on Climate Change. Available from: https://archive.ipcc.ch/ipccreports/tar/wg2/index.php?idp=353 [Accessed on: 2023 Jun 04]

8. Principi N, Campana BR, Argentiero A, Fainardi V, and Esposito S. The Influence of Heat on Pediatric and Perinatal Health: Risks, Evidence, and Future Directions. en. Journal of Clinical Medicine 2025 Feb; 14. doi: 10.3390/jcm14041123. Available from: https://www.mdpi.com/2077-0383/14/4/1123 [Accessed on: 2026 Apr 07]

9. Kioumourtzoglou MA, Raz R, Wilson A, Fluss R, Nirel R, Broday DM, Yuval, Hacker MR, McElrath TF, Grotto I, Koutrakis P, and Weisskopf MG. Traffic-Related Air Pollution and Pregnancy Loss. en. Epidemiology (Cambridge, Mass.) 2019 Jan; 30:4. doi: 10.1097/EDE.0000000000000918. Available from: https://pmc.ncbi.nlm.nih.gov/articles/PMC6269216/ [Accessed on: 2024 Nov 19]

10. Bilancio demografico mensile gennaio-dicembre 2025 – Istat. Available from: https://www.istat.it/notizia/bilancio-demografico-mensile-gennaio-dicembre-2025/ [Accessed on: 2026 Apr 27]

11. EpiCentro. Rapporto sull’evento nascita in Italia (CeDAP) - anno 2022. it. Available from: https://www.epicentro.iss.it/materno/cedap-2022 [Accessed on: 2025 Oct 27]

12. Muñoz-Sabater J, Dutra E, Agustí-Panareda A, Albergel C, Arduini G, Balsamo G, Boussetta S, Choulga M, Harrigan S, Hersbach H, et al. ERA5-Land: A state-of-the-art global reanalysis dataset for land applications. Earth system science data 2021; 13:4349–83

13. Copernicus Atmosphere Monitoring Service. CAMS Global Reanalysis (EAC4). Accessed on. 2025. Available from: https://www.ecmwf.int/en/forecasts/dataset/cams-global-reanalysis

14. Indice di fragilità comunale (IFC). it-IT. Available from: https://www.istat.it/comunicato-stampa/indice-di-fragilita-comunale-ifc/ [Accessed on: 2025 Sep 17]

15. Riebler A, Sørbye SH, Simpson D, and Rue H. An intuitive Bayesian spatial model for disease mapping that accounts for scaling. eng. Statistical Methods in Medical Research 2016 Aug; 25:1145–65. doi: 10.1177/0962280216660421

16. R Core Team. R: A Language and Environment for Statistical Computing. R Foundation for Statistical Computing. Vienna, Austria, 2024. Available from: https://www.R-project.org/

17. R-INLA Project. de. Available from: https://www.r-inla.org/ [Accessed on: 2025 Oct 12]

18. Gasparrini A, Armstrong B, and Scheipl F. Distributed lag linear and non-linear models in R: the package dlnm. Journal of Statistical Software 2011; 43:1–20. doi: 10.18637/jss.v043.i08. Available from: https://doi.org/10.18637/jss.v043.i08

19. Das S, Sagar S, Chowdhury S, Akter K, Haq MZ, and Hanifi SMA. The risk of miscarriage is associated with ambient temperature: evidence from coastal Bangladesh. en. Frontiers in Public Health 2023 Nov; 11:1238275. doi: 10.3389/fpubh.2023.1238275. Available from: https://www.frontiersin.org/articles/10.3389/fpubh.2023.1238275/full [Accessed on: 2024 Nov 19]

20. Wesselink AK, Gause EL, Spangler KR, Hystad P, Kirwa K, Willis MD, Wellenius GA, and Wise LA. Exposure to Ambient Heat and Risk of Spontaneous Abortion: A Case-Crossover Study. eng. Epidemiology (Cambridge, Mass.) 2024 Nov; 35:864–73. doi: 10.1097/EDE.0000000000001774

21. Hajdu T and Hajdu G. Post-conception heat exposure increases clinically unobserved pregnancy losses. eng. Scientific Reports 2021 Jan; 11:1987. doi: 10.1038/s41598-021-81496-x

22. Shah AP, Achilleos S, Wang VA, Leung M, Weisskopf MG, Kyprianou T, Koutrakis P, and Papatheodorou S. Associations of climatic factors with pregnancy loss in Nicosia, Cyprus. eng. Journal of Exposure Science & Environmental Epidemiology 2025 Sep; 35:831–8. doi: 10.1038/s41370-025-00781-3

23. Nanas I, Chouzouris TM, Dovolou E, Dadouli K, Stamperna K, Kateri I, Barbagianni M, and Amiridis GS. Early embryo losses, progesterone and pregnancy associated glycoproteins levels during summer heat stress in dairy cows. Journal of Thermal Biology 2021 May; 98:102951. doi: 10.1016/j.jtherbio.2021.102951. Available from: https://www.sciencedirect.com/science/article/pii/S0306456521001182 [Accessed on: 2025 Oct 30]

24. Muhamad SN, Md Akim A, Lim FL, Karuppiah K, Mohd Shabri NSA, and How V. Heat stress-induced heat shock protein 70 (HSP70) expressions among vulnerable populations in urban and rural areas Klang Valley, Malaysia. en. Journal of Exposure Science & Environmental Epidemiology 2025 Sep; 35:839–47. doi: 10.1038/s41370-025-00764-4. Available from: https://www.nature.com/articles/s41370-025-00764-4 [Accessed on: 2025 Oct 30]

25. Dutta S, Sengupta P, and Liew FF. Cytokine landscapes of pregnancy: mapping gestational immune phases. en. Gynecology and Obstetrics Clinical Medicine 2024 Apr; 4:e000011. doi: 10.1136/gocm-2024-000011. Available from: http://gocm.bmj.com/lookup/doi/ 10.1136/gocm-2024-000011 [Accessed on: 2026 May 11]

26. Alves C, Jenkins SM, and Rapp A. Early Pregnancy Loss (Spontaneous Abortion). eng. StatPearls. Treasure Island (FL): StatPearls Publishing, 2025. Available from: http://www.ncbi.nlm.nih.gov/books/NBK560521/ [Accessed on: 2025 Oct 18]

27. F d, M S, V V, P M, M DS, M D, and P M. Future attributable deaths of heatwaves in Italian cities using high resolution climate change scenarios. en-US. Environmental Epidemiology 2019 Oct; 3:357. doi: 10.1097/01.EE9.0000609956.43849.45. Available from: https://journals.lww.com/environepidem/fulltext/2019/10001/Future_attributable_deaths_of_heatwaves_in_Italian.1092.aspx [Accessed on: 2026 Apr 07]

28. Ministero dell’Ambiente e della Sicurezza Energetica (MASE). Piano Nazionale di Adattamento ai Cambiamenti Climatici (PNACC) [National Climate Change Adaptation Plan]. Tech. rep. Approvato con D.M. n.434 del 21 dicembre 2023 (GU Serie Generale n.42 del 20-02-2024). Accessed: 2026-06-18. Roma: Ministero dell’Ambiente e della Sicurezza Energetica, 2023. Available from: https://www.mase.gov.it/portale/documents/d/guest/pnacc_documento_di_piano-pdf

29. Istituto Superiore di Sanità (ISS). Linea guida Gravidanza Fisiologica [Physiological Pregnancy Clinical Guidelines]. Tech. rep. Accessed: 2026-06-18. Roma: Istituto Superiore di Sanità / Sistema Nazionale Linee Guida (SNLG). Available from: https://snlg.iss.it/

30. Gómez-Rubio V. Bayesian inference with INLA. Chapman and Hall/CRC, 2020

